# The contribution of genetic risk and lifestyle factors in the development of adult-onset inflammatory bowel disease: a prospective cohort study

**DOI:** 10.1101/2022.07.18.22277738

**Authors:** Yuhao Sun, Shuai Yuan, Xuejie Chen, Jing Sun, Rahul Kalla, Lili Yu, Lijuan Wang, Xuan Zhou, Xiangxing Kong, Therese Hesketh, Gwo-tzer Ho, Kefeng Ding, Malcolm Dunlop, Susanna C. Larsson, Jack Satsangi, Jie Chen, Xiaoyan Wang, Xue Li, Evropi Theodoratou, Edward L Giovannucci

## Abstract

**Objective:** There are few prospective data exploring the combined effect of genetic and lifestyle factors on the incidence of inflammatory bowel disease (IBD), and whether genetic risk of IBD may be mitigated by lifestyle remains unclear.

**Design:** We conducted a prospective cohort study based on the UK Biobank to examine the associations across genetic risk, modifiable lifestyle factors, and risk of Crohn’s disease (CD) and ulcerative colitis (UC). Genetic susceptibility to CD and UC was estimated by polygenic risk scores using common genetic variants identified by genome-wide association studies and was further categorized into high, intermediate, and low genetic risk categories. Weighted unhealthy lifestyle scores were constructed based on disease-related lifestyle factors, including ever smoking, unhealthy diet, physical inactivity, obesity, and abnormal sleep duration, and were categorized into ‘favorable’, ‘intermediate’, and ‘unfavorable’ categories. The Cox proportional hazard regression model was used to estimate the HRs and 95% CIs for their associations and accumulative risk for developing CD and UC were estimated for each risk group.

**Results:** During a median follow-up of 12.0 years, 707 CD and 1576 UC cases were diagnosed. Genetic risk and unhealthy lifestyle categories were monotonically associated with CD and UC risk and there was no multiplicative interaction between them. Compared with participants with low genetic risk, the hazard ratios (HRs) of CD and UC were 2.24 (95% confidence interval [CI] 1.75-2.86) and 2.15 (95% CI 1.82-2.53) for those with high genetic risk, respectively. The HRs of CD and UC for individuals in unfavorable lifestyle category were 1.94 (95% CI 1.61-2.33) and 1.98 (95% CI 1.73-2.27), respectively, compared with those in favorable category. When considering genetic risk and lifestyle jointly, the HRs of individuals with high genetic risk but a favorable lifestyle (2.33, 95% CI 1.58-3.44 for CD, and 2.05, 95% CI, 1.58-2.66 for UC) were reduced nearly by half comparing to those with high genetic risk but an unfavorable lifestyle (4.40, 95% CI, 2.91-6.66 for CD and 4.44, 95% CI, 3.34-5.91 for UC).

**Conclusion:** Genetic and lifestyle factors were independently associated with susceptibility to CD and UC. Participants at high genetic risk could reduce nearly 50% risk of CD and UC by adherence to a favorable lifestyle.

**Funding:** XL: the Natural Science Fund for Distinguished Young Scholars of Zhejiang Province (LR22H260001). XYW: National Natural Science Foundation of China (81970494) and Key Project of Research and Development Plan of Hunan Province(2019SK2041); SCL: the Swedish Heart-Lung Foundation (Hjärt-Lungfonden, 20210351), the Swedish Research Council (Vetenskapsrådet, 2019-00977), and the Swedish Cancer Society (Cancerfonden); ET: CRUK Career Development Fellowship (C31250/A22804); KFD: Project of the regional diagnosis and treatment center of the Health Planning Committee (No. JBZX-201903).

**What is already known on this topic:** Previous studies have identified a number of genetic variants and several modifiable risk factors for IBD. However, there is a lack of studies on the combined effects of genetic and lifestyle factors on IBD risk.

**What this study adds:** In this prospective cohort study, genetic risk and modifiable lifestyle factors were independently associated with the risk of incident Crohn’s disease and ulcerative colitis. Participants at high genetic risk could reduce nearly 50% of their genetic susceptibility to Crohn’s disease and ulcerative colitis by adherence to a favorable lifestyle.

**How this study might affect research, practice or policy:** Promoting a healthy lifestyle is an effective strategy to lower the incidence of these diseases, especially among those with high-risk genetic background.

## Introduction

Inflammatory bowel disease (IBD), which includes two main subtypes (Crohn’s disease, CD, and ulcerative colitis, UC), is a global health problem with substantial disease burden, especially in the industrialized countries.^1 2^ Although onset in childhood and early adulthood is well recognized, epidemiological studies now highlight the increasing incidence and prevalence of IBD onset in middle age, or later life. Compared to IBD in children or adolescents, the etiology of adult-onset IBD is believed to be more multifactorial, with genetic and environmental factors playing important roles in its development.^3 4^

Genome-wide association studies have identified over a hundred of risk loci, such as *TYK2, IL2RA*, and *IL23R*, to be associated with IBD.^3 5-9^ Although a single genetic variant accounts for only a small fraction of the genetic variability of IBD, polygenic risk scores combining multiple risk loci can be used as an indicator to identify individuals at higher genetic susceptibility to IBD.^10^ Compared with rare genetic mutations with larger effect (e.g., *NOD2*), polygenic risk scores can identify a larger fraction of population at comparable or greater disease risk, which poses opportunities for clinical utility. However, it is largely unknown whether a PRS of IBD can identify individuals at high genetic risk for potential personalized prevention via adoption of healthy lifestyles in later life.

Observational studies have identified several potentially modifiable risk factors in relation to IBD, including cigarette smoking, unhealthy diet, physical inactivity, obesity, and abnormal sleep duration.^4 11^ The associations of these lifestyle factors with the risk of CD and UC appear complex. For instance, active smoking has been reported to be protective against UC but risky for CD.^11^ The association between alcohol drinking and IBD is inconclusive and remains elusive.^12^ Sleep is one of the common lifestyle factors in maintaining physical and psychological health, however, whether sleep behavior is associated with IBD risk has been scarcely investigated.^13^ Comprehensive appraisal of the associations between these modifiable lifestyle factors and IBD risk will deepen the understanding of the etiology of IBD and provide clues for IBD prevention.

So far, there is lack of comprehensive investigation on the combined effect of genetic and lifestyle factors on the development of IBD and its subtypes. Herein, we conducted a prospective cohort study based on the UK Biobank to examine the associations across genetic risk, modifiable lifestyle factors, and risk of IBD, to test whether there is any multiplicative interaction between genetic risk and lifestyle factors, and to figure out to what extent the genetic risk of IBD may be mitigated by adherence to healthy lifestyle choices.

## Methods

### Study population

This cohort study is based on data collected from the UK Biobank including approximately 500,000 participants recruited across the United Kingdom between 2006 and 2010.^14^ Individuals of non-European ancestry (due to limited numbers and to minimize population structure bias), without genetic information, or with baseline IBD diagnosis, new onset IBD within 1-year follow-up, or unclear IBD diagnosis were excluded, leaving 453,492 individuals (**Figure 1**).

**Figure 1.**
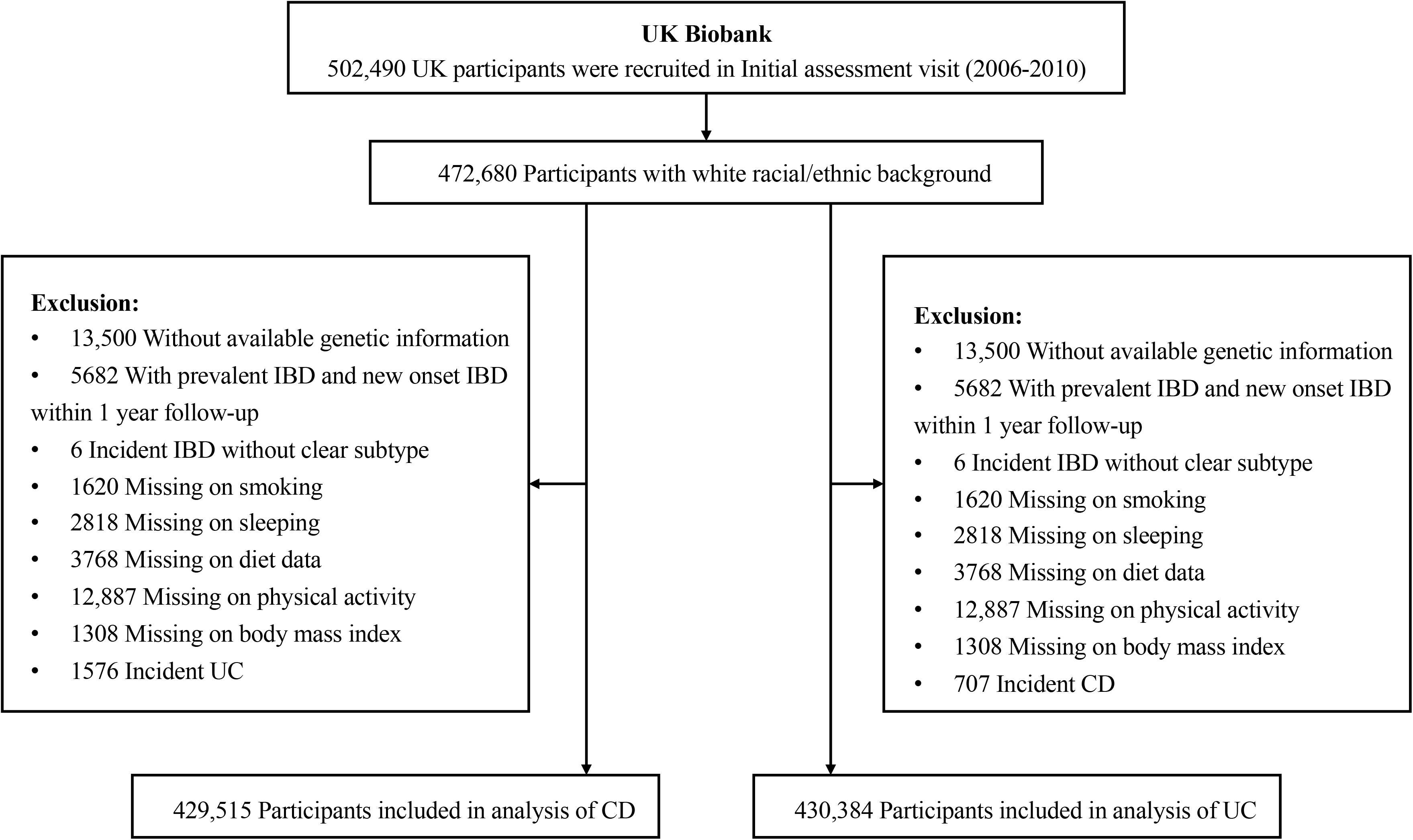
Flowchart of study population selection in the UK Biobank study. CD, Crohn’s disease; IBD, inflammatory bowel disease; UC, ulcerative colitis.

### Genetic risk profiling

We applied two strategies to estimate the genetic susceptibility to CD and UC for UK Biobank population. We firstly constructed a polygenetic risk score (PRS), using the common genetic variants that were identified to be strongly associated with CD and UC (*P*<5×10^−8^) from a genome-wide association meta-analysis of up to 86,640 individuals of European ancestry, including 5,956 CD and 6,968 UC cases.^3^ After removing genetic variants in linkage disequilibrium (*r*^2^>0.001), 51 and 30 independent single nucleotide polymorphisms (SNPs) were used to calculate the PRS of CD and UC, respectively. Polygenic risk scores were constructed for each participant by summing up the number of risk-increasing alleles for each SNP weighted by effect size on genetic liability to CD or UC (**Supplementary Table 1**). Because using merely the genome-wide significant SNPs may omit some correlated informative signals that are independently associated with CD and UC, we additionally constructed genomic risk scores by including all SNPs at suggestive significance level (*P*<1×10^−5^) reported by the GWAS. Genomic risk scores were calculated by using the LDpred2.^15^ Either polygenic risk score or genomic risk score with better stratification ability was taken froward to proxy the genetic susceptibility of CD and UC, and was further used to categorize the low (the lowest quintile), intermediate (quintiles 2 to 4), and high (highest quintile) genetic risk groups.

### Modifiable lifestyle factors

Six common lifestyle factors, including cigarette smoking, diet, alcohol consumption, physical inactivity, BMI, and sleep duration, were examined for their associations with CD and UC risk respectively. These lifestyle factors were chosen based on pre-existing evidence on their associations with either CD or UC, as reported by a recent umbrella review and cohort studies.^11 16^ Detailed information on definitions of common lifestyle factors is displayed in **Supplementary Method and Supplementary Table 2**. Healthy lifestyle scores (HLS) were constructed based on aforementioned lifestyle factors. Individuals were assigned 1 point for each of lifestyle behaviors if they were classified into the healthy group. A higher lifestyle score indicates higher adherence to healthy lifestyle. The unweighted lifestyle score was categorized as favorable (4 or 5 healthy lifestyle factors), intermediate (3 healthy lifestyle factors), and unfavorable (0-2 healthy lifestyle factor) lifestyles. We then constructed a weighted standardized healthy lifestyle score based on the β coefficient of each lifestyle factor in the Cox proportional hazards model adjusted for age, age-square, sex, education, Townsend deprivation index, and first 20 principal components of ancestry using the formula 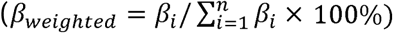 and then the weighted standardized healthy lifestyle score was categorized into unfavorable (the lowest quintile), intermediate (quintiles 2-4), and favorable (the highest quintile) groups.

### Cases ascertainment and follow-up

Diagnostic information was obtained from the primary care data and hospital inpatient records. Incident CD and UC cases were ascertained by a primary or secondary diagnosis defined by corresponding International Classification of Diseases codes (ICD-9: 555, 556; ICD-10: K50, K51). Participants were followed up from the baseline (2006-2010) until the date of first diagnosis of IBD, date of death, date of loss to follow-up, the last date of hospital admission (i.e., HES and SMR: 31 March 2021, and PEDW: 28 February 2018), whichever came first. Disease locations were obtained from diagnosis records for subgroup analyses.^17^

### Statistical analysis

We used the Cox proportional hazard regression model to examine the associations of genetic risk categories, lifestyle categories, and genetic risk and lifestyle combined categories (9 categories with high genetic risk and unfavorable lifestyle as reference) with risk of incident CD and UC. The model was adjusted for age, age-square, sex, education, Townsend deprivation index, Charlson comorbidities index and first 20 principal components of ancestry. The interactions between lifestyle factors and polygenic risk scores were also examined using a multiplicative interaction model. The proportionality of hazards assumption was assessed using the Schoenfeld residuals method and found to be satisfied (*P*>0.15). To examine the consistency of the association in subpopulations, we conducted stratification analyses by age (≥60 and <60 years), sex (female and male), education attainment (≥college/university and < college/university), and the tertiles of Townsend deprivation index (from low to high, T1-3). We also stratified the analysis on the associations of the healthy lifestyle categories with CD and UC risk by genetic risk. Sensitivity analyses and subgroup analyses by considering disease locations and age of diagnosis of UC and CD (to retrospectively include prevalent cases and stratify analysis by age of onset) were also performed to thoroughly examine their complex associations (**Supplementary Method**). The cumulative incidence of CD and UC by categories of genetic risk and lifestyle scores were obtained using the cumulative incidence function of competing risk regression.^18^ All tests were two-sided and the association with the *P* value <0.05 was deemed significant. All analyses were performed using R software, version 3.6.3.

## Results

**Table 1** shows baseline characteristics of included participants by incident disease status. Over a median follow-up of 12.0 years (interquartile range, 11.2-12.7 years), 707 CD and 1576 UC cases were diagnosed. The mean age of diagnosis was 64 (range: 43-82 for CD and 65 (range: 43-82) for UC.

**Table 1.** Characteristics of participants by incident Crohn’s disease and ulcerative colitis status.

| Characteristic | Participants included in the analyses of Crohn's disease |  |  |  | Participants included in the analyses of ulcerative colitis |  |  |  |
| --- | --- | --- | --- | --- | --- | --- | --- | --- |
|  | Incident Crohn's disease status |  |  | All participants (n=429,515) | Incident ulcerative colitis status |  |  | All participants (n=430,384) |
|  | No (n=428,808) | Yes (n=707) | P value |  | No (n=428,808) | Yes (n=1576) | P value |  |
| Age, mean (SD) | 56.7 (8.0) | 57.1 (8.2) | 0.187 | 56.7 (8.0) | 56.7 (8.0) | 57.7 (7.8) | <0.001 | 56.7 (8.0) |
| Female | 233,956 (54.6) | 402 (56.9) | 0.234 | 234,358 (54.6) | 233,956 (54.6) | 753 (47.8) | <0.001 | 234,709 (54.5) |
| TDI, mean (SD) | -1.5 (3.0) | -0.9 (3.2) | <0.001 | -1.5 (3.0) | -1.5 (3.0) | -1.1 (3.2) | <0.001 | -1.5 (3.0) |
| College/University degree | 141862 (33.1) | 186 (26.3) | <0.001 | 142048 (33.1) | 141862 (33.1) | 389 (24.7) | <0.001 | 142251 (33.1) |
| CCI, mean (SD) | 0.3 (0.9) | 0.4 (1.0) | <0.001 | 0.3 (0.9) | 0.3 (0.9) | 0.4 (1.0) | <0.001 | 0.3 (0.9) |
| Healthy lifestyle factors |  |  |  |  |  |  |  |  |
| Never smoking | 233,201 (54.4) | 324 (45.8) | <0.001 | 233,525 (54.4) | 233,201 (54.4) | 634 (40.2) | <0.001 | 233,835 (54.3) |
| No obesity | 326,755 (76.2) | 483 (68.3) | <0.001 | 327,238 (76.2) | 326,755 (76.2) | 1146 (72.7) | 0.001 | 327,901 (76.2) |
| Adequate sleep duration (7-8h) | 294,953 (68.8) | 435 (61.5) | <0.001 | 295,388 (68.8) | 294,953 (68.8) | 994 (63.1) | <0.001 | 295,947 (68.8) |
| Healthy diet | 297,426 (69.4) | 451 (63.8) | 0.002 | 297,877 (69.4) | 297,426 (69.4) | 1004 (63.7) | <0.001 | 298,430 (69.3) |
| Regular physical activity | 336,897 (78.6) | 517 (73.1) | 0.001 | 337,414 (78.6) | 336,897 (78.6) | 1189 (75.4) | 0.003 | 338,086 (78.6) |
| Number of healthy lifestyle factors |  |  | <0.001 |  |  |  | <0.001 |  |
| 0 | 2878 (0.7) | 11 (1.6) |  | 2889 (0.7) | 2878 (0.7) | 15 (1.0) |  | 2893 (0.7) |
| 1 | 19,035 (4.4) | 57 (8.1) |  | 19,092 (4.4) | 19,035 (4.4) | 117 (7.4) |  | 19,152 (4.4) |
| 2 | 59,861 (14.0) | 144 (20.4) |  | 60,005 (14.0) | 59,861 (14.0) | 321 (20.4) |  | 60,182 (14.0) |
| 3 | 119,137 (27.8) | 209 (29.6) |  | 119,346 (27.8) | 119,137 (27.8) | 482 (30.6) |  | 119,619 (27.8) |
| 4 | 146,421 (34.1) | 192 (27.2) |  | 146,613 (34.1) | 146,421 (34.1) | 443 (28.1) |  | 146,864 (34.1) |
| 5 | 81,476 (19.0) | 94 (13.3) |  | 81,570 (19.0) | 81,476 (19.0) | 198 (12.6) |  | 81,674 (19.0) |
| Genetic risk category <sup>a</sup> |  |  | <0.001 |  |  |  | <0.001 |  |
| Low | 85,811 (20.0) | 92 (13.0) |  | 85,903 (20.0) | 85,865 (20.0) | 212 (13.5) |  | 86,077 (20.0) |
| Intermediate | 257,301 (60.0) | 408 (57.7) |  | 257,709 (60.0) | 257,320 (60.0) | 910 (57.7) |  | 258,230 (60.0) |
| High | 85,696 (20.0) | 207 (29.3) |  | 85,903 (20.0) | 85,623 (20.0) | 454 (28.8) |  | 86,077 (20.0) |

TDI indicates Townsend deprivation index; CCI, Charlson comorbidity index.
<sup>a</sup> Genetic risk category was defined as low (the lowest quintile), intermediate (2-4 quintiles), and high (the highest quintile) according to polygenic risk scores.

For genetic susceptibility, both polygenic risk score and genomic risk score showed significantly associations with the risk of CD and UC (**Supplementary Table 4**). Comparing to the PRS, the genomic risk score showed no further improvement on the stratification of genetic risk groups (**Table 2, Supplementary Table 4**); therefore, only the PRS was used in the joint analysis. The PRS was normally distributed (**Supplementary Figure 1**) and showed no associations with lifestyle factors with the exception for an association between polygenic risk score of CD and smoking status (**Supplementary Table 5**). Risk of incident CD and UC increased across genetic risk categories (low to high) in a linear fashion (**Table 2**). Compared with participants with low genetic risk, the hazard ratios (HRs) of CD and UC were 2.24 (95% confidence interval, CI: 1.75-2.86, *P*<0.001) and 2.15 (95% CI: 1.82-2.53, *P*<0.001), for those with high genetic risk, respectively. The associations remained significant after additional adjustment for lifestyle factors. The same pattern of associations was observed in the analysis using the genetic risk quintiles instead of categories (**Supplementary Table 4**).

**Table 2.**
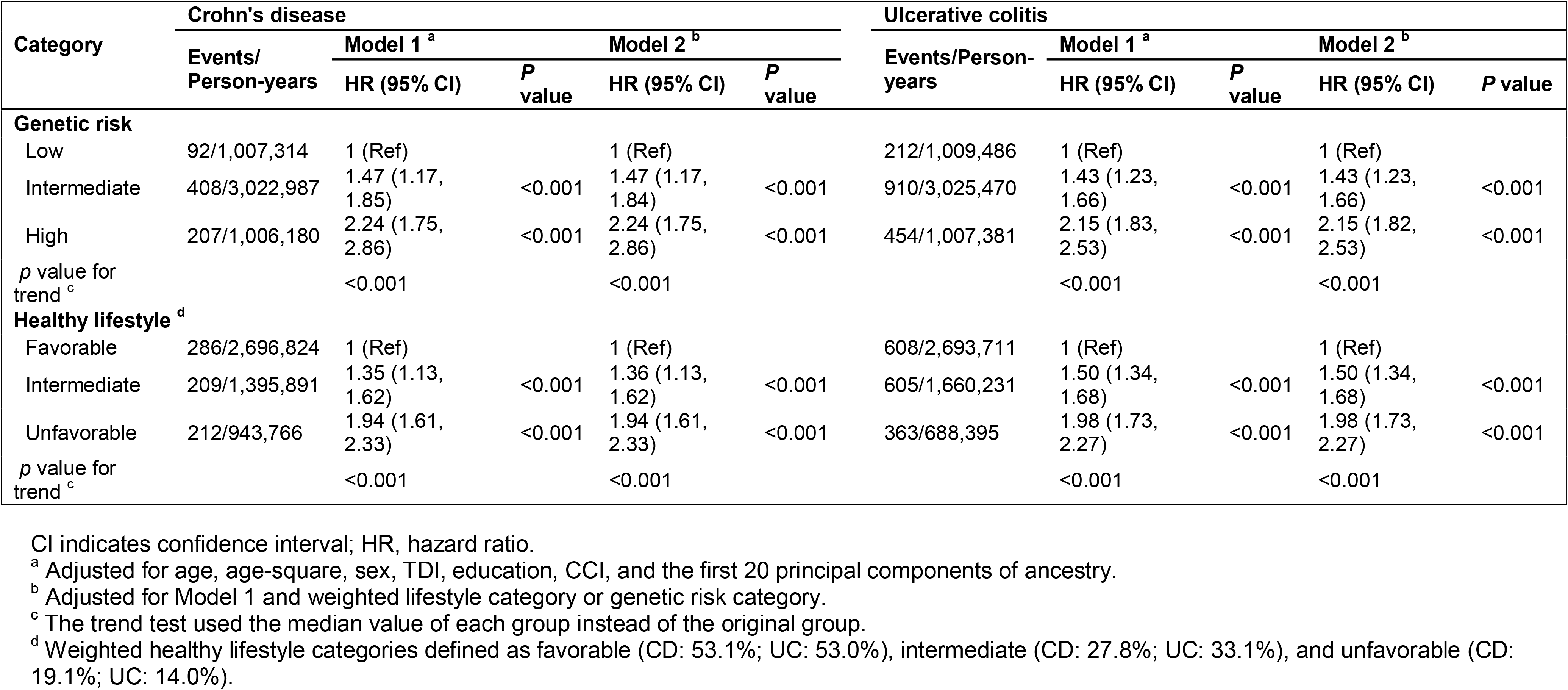
Risk of incident Crohn’s disease and ulcerative colitis according to genetic risk and lifestyle categories.

| Category | Crohn's disease |  |  |  |  | Ulcerative colitis |  |  |  |  |
| --- | --- | --- | --- | --- | --- | --- | --- | --- | --- | --- |
|  | Events/<br>Person-years | Model 1 <sup>a</sup> |  | Model 2 <sup>b</sup> |  | Events/Person-years | Model 1 <sup>a</sup> |  | Model 2 <sup>b</sup> |  |
|  |  | HR (95% CI) | *P*<br>value | HR (95% CI) | *P*<br>value |  | HR (95% CI) | *P*<br>value | HR (95% CI) | *P*<br>value |
| **Genetic risk** |  |  |  |  |  |  |  |  |  |  |
| Low | 92/1,007,314 | 1 (Ref) |  | 1 (Ref) |  | 212/1,009,486 | 1 (Ref) |  | 1 (Ref) |  |
| Intermediate | 408/3,022,987 | 1.47 (1.17, 1.85) | <0.001 | 1.47 (1.17, 1.84) | <0.001 | 910/3,025,470 | 1.43 (1.23, 1.66) | <0.001 | 1.43 (1.23, 1.66) | <0.001 |
| High | 207/1,006,180 | 2.24 (1.75, 2.86) | <0.001 | 2.24 (1.75, 2.86) | <0.001 | 454/1,007,381 | 2.15 (1.83, 2.53) | <0.001 | 2.15 (1.82, 2.53) | <0.001 |
| *p* value for trend <sup>c</sup> |  | <0.001 |  | <0.001 |  |  | <0.001 |  | <0.001 |  |
| **Healthy lifestyle <sup>d</sup>** |  |  |  |  |  |  |  |  |  |  |
| Favorable | 286/2,696,824 | 1 (Ref) |  | 1 (Ref) |  | 608/2,693,711 | 1 (Ref) |  | 1 (Ref) |  |
| Intermediate | 209/1,395,891 | 1.35 (1.13, 1.62) | <0.001 | 1.36 (1.13, 1.62) | <0.001 | 605/1,660,231 | 1.50 (1.34, 1.68) | <0.001 | 1.50 (1.34, 1.68) | <0.001 |
| Unfavorable | 212/943,766 | 1.94 (1.61, 2.33) | <0.001 | 1.94 (1.61, 2.33) | <0.001 | 363/688,395 | 1.98 (1.73, 2.27) | <0.001 | 1.98 (1.73, 2.27) | <0.001 |
| *p* value for trend <sup>c</sup> |  | <0.001 |  | <0.001 |  |  | <0.001 |  | <0.001 |  |

CI indicates confidence interval; HR, hazard ratio.
<sup>a</sup> Adjusted for age, age-square, sex, TDI, education, CCI, and the first 20 principal components of ancestry.
<sup>b</sup> Adjusted for Model 1 and weighted lifestyle category or genetic risk category.
<sup>c</sup> The trend test used the median value of each group instead of the original group.
<sup>d</sup> Weighted healthy lifestyle categories defined as favorable (CD: 53.1%; UC: 53.0%), intermediate (CD: 27.8%; UC: 33.1%), and unfavorable (CD: 19.1%; UC: 14.0%).

The associations of individual lifestyle factors with the risk of CD and UC are presented in **Table 3**. Less healthy behavior was in general associated with increased risk of CD and UC, compared with those meeting healthy lifestyle guidelines (the reference category) for each component of healthy lifestyles although not all risk estimates were statistically significant. Exceptions were noted for alcohol drinking, which was neither associated with CD nor associated with UC (**Supplementary Table 6**). Given that there was no well-established evidence supporting their associations from previous evidence either,^12^ we therefore excluded alcohol consumption from the construction of healthy lifestyle scores. For UC, the association with obesity was not statistically significant (*P*=0.29) and irregular physical activity was marginally associated with an increased risk of UC (*P*=0.068); nevertheless, given that obesity and physical activity were well-established lifestyle factors related to IBD based on previous evidence,^19 20^ we decided to include these variables for the construction of healthy lifestyle scores. The associations of smoking status and frequency with the risk of CD and UC at different disease locations and age of diagnosis are shown with details in **Supplementary Tables 7-11**. Briefly, both previous and current smoking were consistently associated with an increased risk of CD and UC among older populations (**Supplementary Table 11)**. We therefore simplified the smoking exposure into ever versus never smoking behaviors as one of the components of a healthier lifestyle.

**Table 3.** Risk of incident Crohn’s disease and ulcerative colitis with each lifestyle factor

| Lifestyle Factors | Incident CD |  |  |  | Incident UC |  |  |  |
| --- | --- | --- | --- | --- | --- | --- | --- | --- |
|  | PAR | Events/Person-years | HR (95% CI) | P value | PAR | Events/Person-years | HR (95% CI) | P value |
| Never smoking |  | 324/2,760,887 | 1 (Ref) |  |  | 634/2,763,009 | 1 (Ref) |  |
| Ever smoking | 12.74 | 383/2,275,594 | 1.32 (1.13, 1.53) | <0.001 | 21.79 | 942/2,279,328 | 1.61 (1.45, 1.78) | <0.001 |
| No obesity |  | 483/3,847,349 | 1 (Ref) |  |  | 1146/3,851,809 | 1 (Ref) |  |
| Obesity | 7.29 | 224/1,189,132 | 1.33 (1.13, 1.56) | 0.001 | 1.41 | 430/1,190,528 | 1.06 (0.95, 1.19) | 0.291 |
| Adequate sleep duration (7-8h) |  | 435/3,474,632 | 1 (Ref) |  |  | 994/3,478,454 | 1 (Ref) |  |
| Abnormal sleep duration | 7.51 | 272/1,561,849 | 1.26 (1.08, 1.47) | 0.003 | 5.60 | 582/1,563,883 | 1.19 (1.07, 1.32) | 0.001 |
| Healthy diet |  | 451/3,500,873 | 1 (Ref) |  |  | 1004/3,504,562 | 1 (Ref) |  |
| Unhealthy diet | 6.32 | 256/1,535,608 | 1.22 (1.04, 1.43) | 0.016 | 4.68 | 572/1,537,776 | 1.16 (1.04, 1.29) | 0.006 |
| Regular physical activity |  | 517/3,963,828 | 1 (Ref) |  |  | 1189/3,968,374 | 1 (Ref) |  |
| Irregular physical activity | 4.50 | 190/1,072,653 | 1.22 (1.03, 1.44) | 0.022 | 2.30 | 387/1,073,963 | 1.11 (0.99, 1.25) | 0.068 |

CI indicates confidence interval; HR, hazard ratio; PAR, population attributable risk.
Adjusted for age, age-square, sex, TDI, education, CCI, polygenic risk score of CD or UC, and first 20 principal components of ancestry, and other lifestyle factors.

Having a healthier lifestyle score was significantly related to a reduced risk of CD and UC in a dose-response manner (*P* for trend < 0.001) (**Table 2**). The HRs of CD and UC for individuals in the unfavorable category were 1.94 (95% CI: 1.61-2.33; *P*<0.001) and 1.98 (95% CI: 1.73-2.27; *P*<0.001), respectively, compared with those in the favorable category. The associations did not change in the sensitivity analysis with further adjustment for genetic risk (**Table 2**), in the analysis using the number of healthy lifestyle factors instead of categories (**Supplementary Table 12**) and in the analysis using the unweighted healthy lifestyle score (**Supplementary Table 13**). The cumulative incidence rate of CD and UC during the follow-up was higher in the group with an unfavorable lifestyle compared to the group with a favorable lifestyle (**Figure 2**).

**Figure 2.**
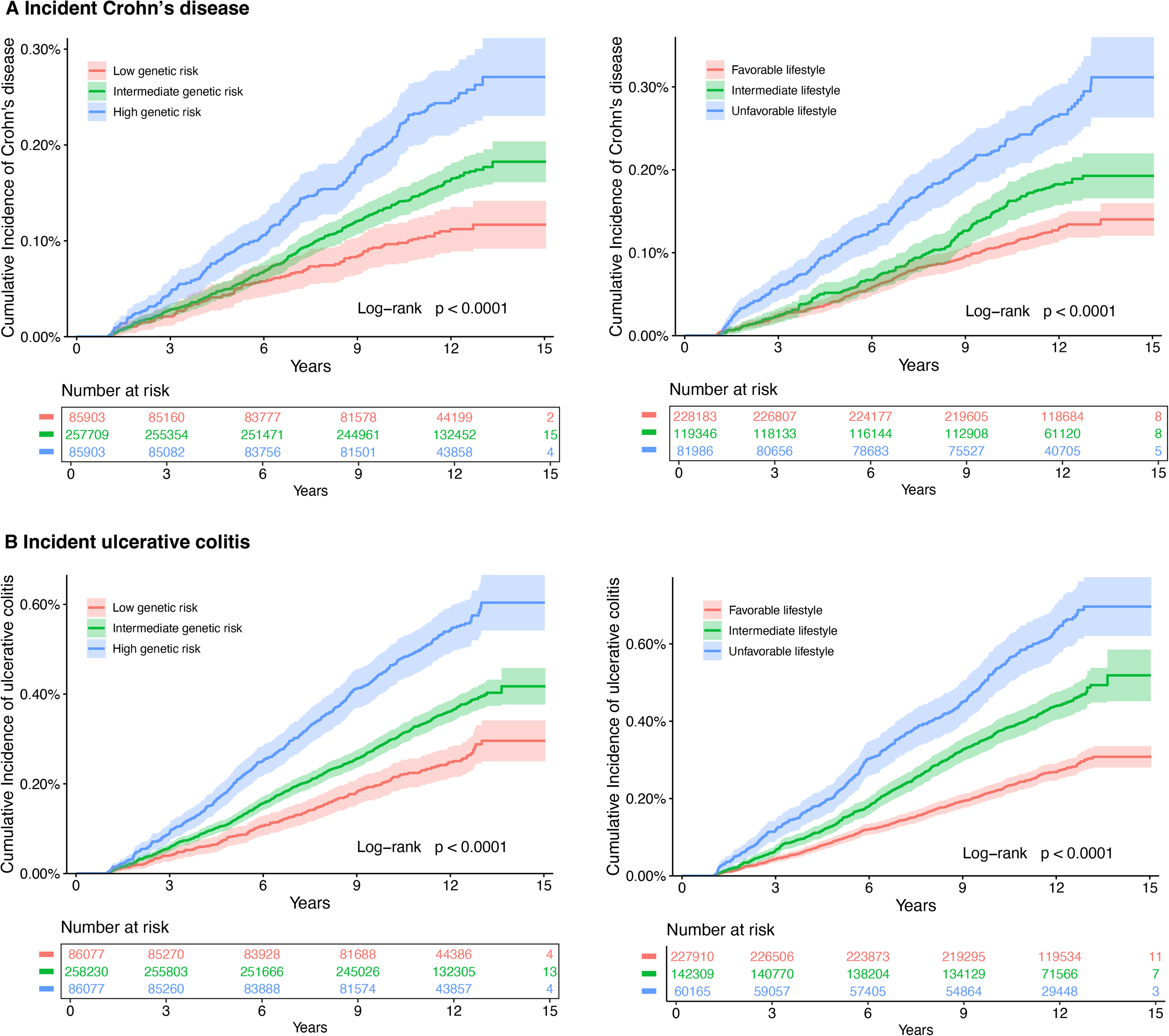
Cumulative incidence plot of the risk of incident Crohn’s disease (A) and ulcerative colitis (B) by polygenic risk score categories and healthy lifestyle categories.

In the analysis of joint categories for genetic risk and healthy lifestyle, the HR of CD and UC showed a linear increase with increasing genetic risk and decreasing healthy lifestyle score (**Figure 3**). Compared with individuals with low genetic risk and favorable lifestyle, the HRs of CD and UC for those with high genetic risk and unfavorable lifestyle were 4.40 (95% CI: 2.91-6.66; *P*<0.001) and 4.44 (95% CI: 3.34-5.91; *P*<0.001), respectively. We observed no significant difference in the HRs of CD or UC between the group of high-genetic risk but having a favorable lifestyle (HR=2.33, 95%CI: 1.58-3.44 for CD and HR=2.05, 95%CI: 1.58-2.66 for UC) and the group of low-genetic risk but having an unfavorable lifestyle (HR=2.32, 95%CI: 1.44-3.74 for CD and HR=1.77, 95%CI: 1.23-2.56 for UC). The analysis on the associations of healthy lifestyle categories with incident CD and UC risk in groups defined by genetic risk confirmed that the unfavorable lifestyle was associated with higher risk of CD and UC across all genetic groups (**Table 4**). Specifically, in individuals with low genetic risk, the HRs of CD and UC were 2.32 (95% CI, 1.42-3.81) and 1.70 (95% CI, 1.17-2.47) for participants with an unfavorable lifestyle compared to those with a favorable lifestyle. We did not detect any multiplicative interaction between the genetic risk and the weighted healthy lifestyle score (*p*=0.85 for CD and *p*=0.87 for UC, **Supplementary Table 14**). The observed associations remained statistically significant in a series of sensitivity analyses (**Supplementary Tables 15**-**20**).

**Table 4.** Risk of incident Crohn’s disease and ulcerative colitis according to lifestyle categories within each genetic risk category.

| Subgroup | Crohn's disease |  |  | Ulcerative colitis |  |  |
| --- | --- | --- | --- | --- | --- | --- |
|  | Events/Person-years | HR (95% CI) | P value | Events/Person-years | HR (95% CI) | P value |
| **Low genetic risk** |  |  |  |  |  |  |
| Favorable lifestyle | 36/537,238 | 1 (Ref) |  | 84/538,409 | 1 (Ref) |  |
| Intermediate lifestyle | 24/281,312 | 1.22 (0.73, 2.06) | 0.452 | 83/332,770 | 1.47 (1.08, 2.00) | 0.014 |
| Unfavorable lifestyle | 32/188,765 | 2.32 (1.42, 3.81) | 0.001 | 45/138,308 | 1.70 (1.17, 2.47) | 0.005 |
| p value for trend<sup>a</sup> |  |  | 0.002 |  |  | 0.002 |
| **Intermediate genetic risk** |  |  |  |  |  |  |
| Favorable lifestyle | 165/1,620,117 | 1 (Ref) |  | 352/1,617,488 | 1 (Ref) |  |
| Intermediate lifestyle | 124/836,165 | 1.41 (1.12, 1.79) | 0.004 | 353/996,460 | 1.52 (1.31, 1.77) | <0.001 |
| Unfavorable lifestyle | 119/566,705 | 1.94 (1.52, 2.47) | <0.001 | 205/411,522 | 1.98 (1.66, 2.37) | <0.001 |
| p value for trend<sup>a</sup> |  |  | <0.001 |  |  |  |
| **High genetic risk** |  |  |  |  |  |  |
| Favorable lifestyle | 85/539,469 | 1 (Ref) |  | 172/537,814 | 1 (Ref) |  |
| Intermediate lifestyle | 61/278,415 | 1.30 (0.94, 1.82) | 0.118 | 169/331,001 | 1.48 (1.20, 1.84) | <0.001 |
| Unfavorable lifestyle | 61/188,296 | 1.80 (1.28, 2.53) | 0.001 | 113/138,566 | 2.13 (1.66, 2.72) | <0.001 |
| p value for trend<sup>a</sup> |  |  | <0.001 |  |  | <0.001 |

CI indicates confidence interval; HR, hazard ratio.
<sup>a</sup> The trend test used the median value of each group instead of the original group.
The analysis was based on weighted healthy lifestyle categories.

**Figure 3.**
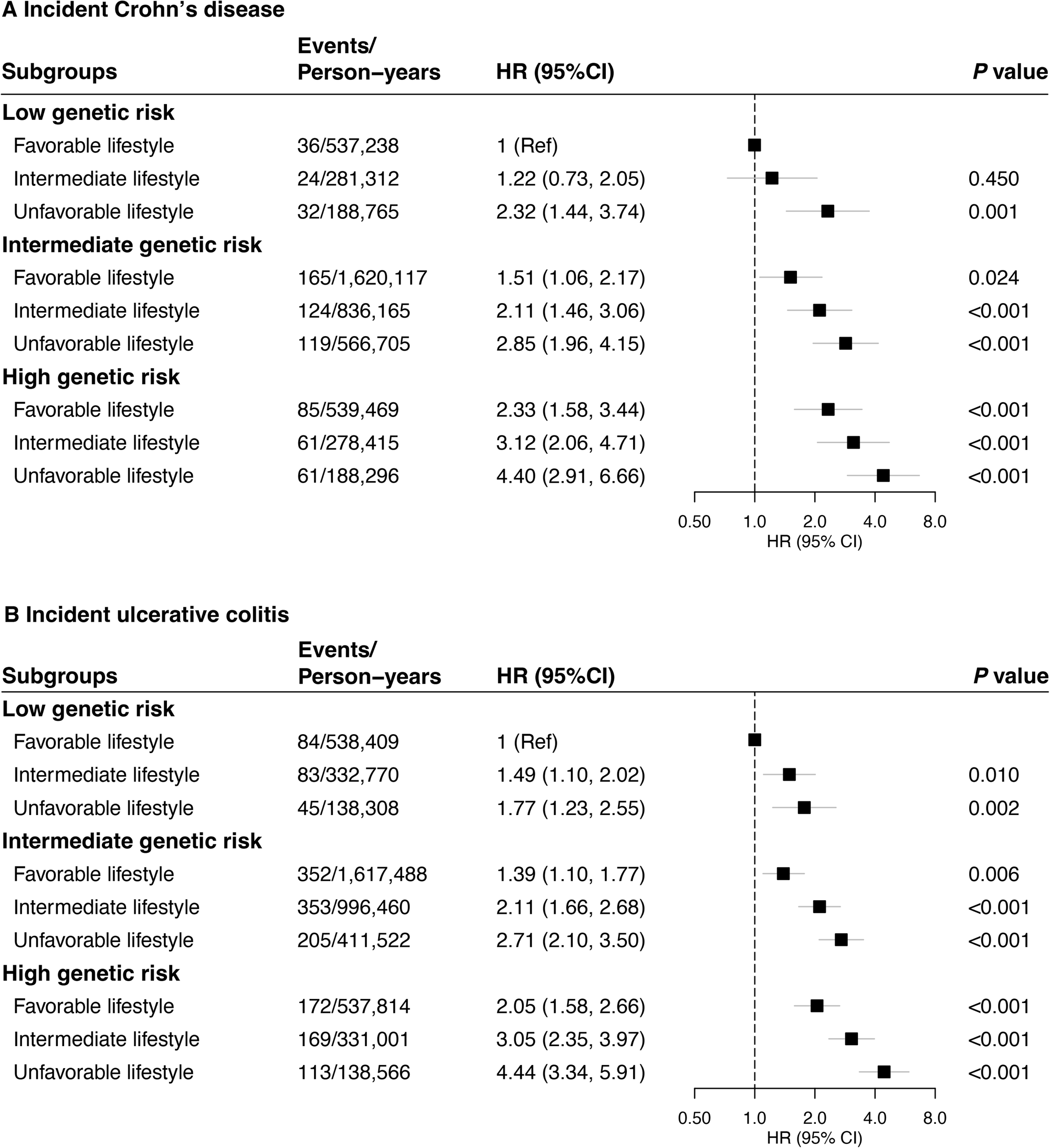
Risk of incident Crohn’s disease and ulcerative colitis by joint categorization for genetic risk and healthy lifestyle score. CI, confidence interval; HR, hazard ratio.

We calculated the cumulative risk of CD and UC over 12 years for each group defined jointly by genetic risk and healthy lifestyle scores (**Figure 4**). Compared to those with low genetic risk and favorable lifestyle (accumulative risk: 0.08% for CD, 0.18% for UC), individuals with high genetic risk and unfavorable lifestyle had 4.87 times higher accumulative risk of CD (equivalent to an excess risk of 0.31% due to high genetic susceptibility and unfavorable lifestyle together) and 5.28 times higher accumulative risk of UC (equivalent to an excess risk of 0.77% due to high genetic susceptibility and unfavorable lifestyle together).

**Figure 4.**
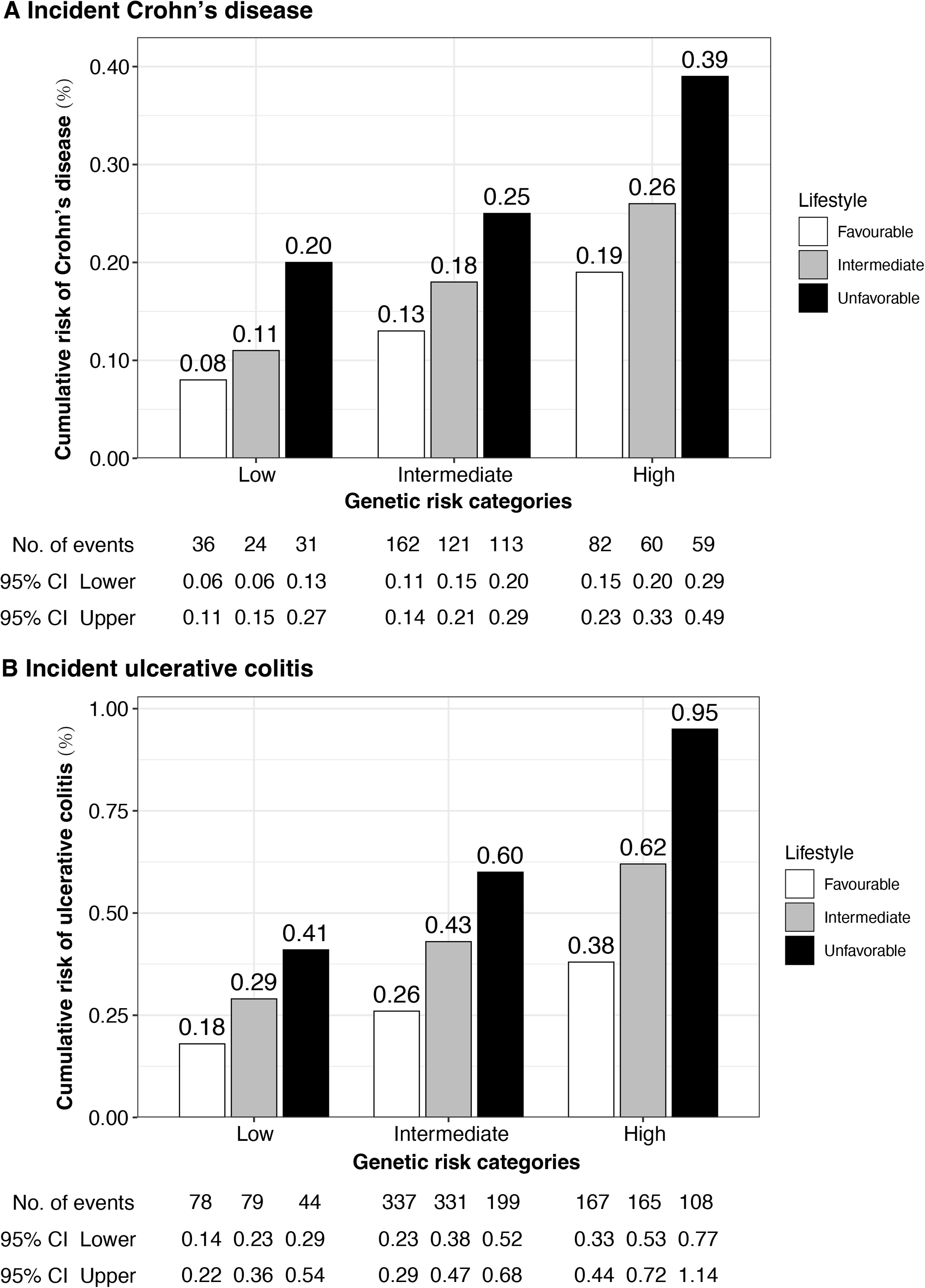
Cumulative incidence plot of the risk of incident Crohn’s disease (A) and ulcerative colitis (B) by joint categorization for genetic risk and healthy lifestyle. CI, confidence interval.

## Discussion

We conducted a cohort study using data from the UK Biobank to investigate the associations across genetic susceptibility, modifiable lifestyle factors, and risk of CD and UC. We found that a polygenic risk score and modifiable lifestyle factors were independently associated with risk of incident CD and UC. No multiplicative interaction was observed between the polygenic risk score and lifestyle scores. Individuals with high genetic risk and an unfavorable lifestyle had elevated risk of CD and UC compared to their counterparts with low genetic risk and a favorable lifestyle. The cumulative incidence of CD and UC over 12 years was approximately five times higher in those with highest genetic risk and unfavorable lifestyle compared to those with the lowest genetic risk and favorable lifestyle.

Even though no studies have been conducted to examine the effect of overall healthy lifestyle on CD or UC risk, a high adherence to a healthy lifestyle was associated with reduced mortality in CD or UC patients from three large cohort studies.^21^ For individual lifestyle factors, there are epidemiological studies assessing the associations of each lifestyle factor with CD and UC risk. Current smoking was found to be positively associated with risk of CD, but inversely associated with risk of UC in a meta-analysis of 9 and 13 studies, respectively.^22^ However, most included studies are cross-sectional studies where residual confounding and reverse causality could not be eliminated. In a subsequent prospective cohort study using data from Nurses’ Health Study (NHS) and Nurses’ Health Study II, the risk of CD was observed to increase in former and current smokers after an over 18 years’ follow-up period.^23^ A positive association between smoking and UC risk was observed in some subsequent studies.^24 25^ Smoking has been also associated with progression of CD, ^26^ but not with that of UC.^27^ In this study, we performed a series of sensitivity/subgroup analyses by considering disease locations and age of diagnosis to thoroughly examine their complex associations with smoking. Our study verified that both previous or current smoking were consistently associated with increased risk of CD regardless of disease locations and age of onset. However, the association between smoking and UC appears complex, in which smoking was associated with reduced risk for early-onset UC (<=20 years) but increased risk for later-onset UC (>40 years). Our previous study showed that smoking habit influences the age at diagnosis and changes in disease extent in UC ^28^. The mechanisms of the observed inverse association are not clear, but reverse causality is again an important point to consider. Since the initial age structure of UK biobank cohort is older, we therefore concentrated on older onset cases and considered smoking as a risk factor of CD and UC.

Evidence on the association between obesity and CD is inconsistent with a positive association in a meta-analysis of 5 cohort studies^20^ but a null finding in another meta-analysis^29^. In a recent prospective analysis of 5 cohorts, obesity defined by BMI was associated with an increased risk of older-onset CD but not UC.^30^ A recent Mendelian randomization study found that genetically predicted higher BMI and body fat percentage was associated with an increased risk of CD, but that genetically predicted higher BMI was associated with a lower risk of UC.^31^ Regular physical activity has been associated with a lower risk of CD, but not UC.^19 32^ Even though a high adherence to Mediterranean diet has associated with a low risk of CD ^33^ and abnormal sleep duration has been associated with a high risk of UC,^34^ there are few corresponding prior studies that examined these associations jointly. Our analysis using data from UK Biobank verified the associations of smoking, physical inactivity, unhealthy diet, and abnormal sleep duration with CD risk, with the exception of alcohol consumption. As for UC, this cohort study found positive associations between all aforementioned lifestyle factors with risk of UC, but no significant association for alcohol drinking either. The null association of alcohol drinking with the risk of CD and UC reported by the present study is consistent with evidence from a recent cohort study.^12^

To our knowledge, no previous studies have examined the association of a combination of healthy lifestyle and multiple genetic factors with risk of incident CD and UC. However, the interaction effects between genes and environmental factors, such as smoking and certain dietary nutrients, on CD and UC have been assessed. A study including 19,735 IBD cases (10,856 CD cases and 8879 UC cases) of known smoking status found that two variants in *HLA* and *NOD2* gene regions interacted with smoking in influencing CD risk and smoking modified the disease risk of some variants in opposite directions for CD vs UC,^35^ which indicated that the effects of smoking on IBD risk depend on genetic variants. Nevertheless, an increased risk of CD and a decreased risk of UC were found in smokers in twin or sibling studies where the cases and controls shared genetic risk for the disease ^36 37^. The interaction effects were also observed for certain dietary nutrients, such as dietary fatty acids, potassium, and iron intake; however, these findings are far from being established to determine the gene-environment interaction on CD and UC risk.^38^ Our study found independent associations of genetic risk and healthy lifestyle with IBD risk, but no overall interaction for their joint effects on IBD risk. Among individuals with high genetic risk, those with unfavorable lifestyle had double the risk of CD or UC compared to those with favorable lifestyle. This finding has important clinical implications by indicating that promoting a healthy lifestyle is an effective strategy to lower incidence of these diseases, even among those with high-risk genetic background.

The strengths of this study include the joint analysis of the genetic and lifestyle factors to gain a comprehensive understanding on the risk of CD and UC, in which polygenetic risk scores and healthy lifestyle scores were constructed to examine their associations with the disease risk in a large prospective cohort of UK Biobank participants.^3^ We made efforts to account for additional genetic susceptibility that were not captured by genome-wide significant SNPs, while the more sophisticated genomic risk score showed no superior capacity comparing to the simple PRS in risk stratification. Limitations of the present study should also be acknowledged when interpreting the findings. Firstly, adherence to a healthy lifestyle might change in the follow-up and influence the association estimation. Nonetheless, the bias caused by the change should be non-differential and therefore attenuate the estimates in a conservative way due to the prospective nature of the design. Secondly, even though important known confounders were adjusted in the models, there is possible residual confounding. Thus, the causality of the association for lifestyle factors cannot be exclusively determined. Thirdly, information on lifestyle factors was partially collected via a self-administrative questionnaire survey, and thus the probability of misclassifications of lifestyle factors and recall bias cannot be fully excluded. This non-differential bias is likely to drive these findings toward to null. Additionally, misclassification of outcome possibly caused by cases undocumented in medical records could attenuate the effect estimates. Fourthly, given that the current analyses were confined to individuals of European ancestry and concentrated more on older population, our findings may not be generalizable to other populations of different ethnicities and/ or ages.

In summary, both high genetic risk and an unfavorable lifestyle were associated with increased risk of CD and UC among adults without IBD. An unfavorable lifestyle was associated with higher risk of CD and UC in individuals regardless of genetic strata. Participants at high genetic risk could reduce nearly 50% risk of CD and UC by adherence to a favorable lifestyle.

## Data Availability

Researchers can request the data we used from the UK Biobank (www.ukbiobank.ac.uk/).

## Additional information

## Acknowledgement

This research was conducted using the UK Biobank study under Application Number 66354. We want to thank all UK Biobank participants and the management team for their participation and assistance.

## Contributors

XL, JC and XYW conceptualized the project. YHS, SY and XJC performed the analyses and wrote the first draft. JS, LLY, LJW, XXK, TH, XK, GH, MD, KFD, SCL, JS, ET and ELG helped with the review and editing. XL, JC and XYW revised the final version. XL, JC and XYW contributed equally to this work and should be considered co-corresponding authors. All authors critically reviewed the manuscript for important intellectual content. XL is the study guarantor. The corresponding author attests that all listed authors meet authorship criteria and that no others meeting the criteria have been omitted.

## No competing interests

“All authors have completed the ICMJE uniform disclosure form at http://www.icmje.org/disclosure-of-interest/ and declare: no support from any organization for the submitted work; no financial relationships with any organizations that might have an interest in the submitted work in the previous three years; no other relationships or activities that could appear to have influenced the submitted work.”

## Ethical approval

The manuscript didn’t previously publish or be under consideration for publication elsewhere. And the ethical approval was granted for the UK Biobank by the North West-Haydock Research Ethics Committee (REC reference: 16/NW/0274).

## Data sharing

Researchers can request the data we used from the UK Biobank (www.ukbiobank.ac.uk/).

## Transparency

The lead author (XL) affirms that the manuscript is an honest, accurate, and transparent account of the study being reported; that no important aspects of the study have been omitted; and that any discrepancies from the study as planned have been explained.

## Dissemination to participants and related patient and public communities

The results of the research will be disseminated to the public through broadcasts, popular science articles, and newspapers.

## References

1. Ng SC, Shi HY, Hamidi N, et al. Worldwide incidence and prevalence of inflammatory bowel disease in the 21st century: a systematic review of population-based studies. Lancet 2017;390(10114):2769–78. doi: 10.1016/s0140-6736(17)32448-0 [published Online First: 2017/10/21]

2. Jairath V, Feagan BG. Global burden of inflammatory bowel disease. Lancet Gastroenterol Hepatol 2020;5(1):2–3. doi: 10.1016/s2468-1253(19)30358-9 [published Online First: 2019/10/28]

3. Liu JZ, van Sommeren S, Huang H, et al. Association analyses identify 38 susceptibility loci for inflammatory bowel disease and highlight shared genetic risk across populations. Nat Genet 2015;47(9):979–86. doi: 10.1038/ng.3359 [published Online First: 2015/07/21]

4. Rozich JJ, Holmer A, Singh S. Effect of Lifestyle Factors on Outcomes in Patients With Inflammatory Bowel Diseases. Am J Gastroenterol 2020;115(6):832–40. doi: 10.14309/ajg.0000000000000608 [published Online First: 2020/04/01]

5. Huang H, Fang M, Jostins L, et al. Fine-mapping inflammatory bowel disease loci to single-variant resolution. Nature 2017;547(7662):173–78. doi: 10.1038/nature22969 [published Online First: 2017/06/29]

6. de Lange KM, Moutsianas L, Lee JC, et al. Genome-wide association study implicates immune activation of multiple integrin genes in inflammatory bowel disease. Nat Genet 2017;49(2):256–61. doi: 10.1038/ng.3760 [published Online First: 2017/01/10]

7. Hugot JP, Chamaillard M, Zouali H, et al. Association of NOD2 leucine-rich repeat variants with susceptibility to Crohn’s disease. Nature 2001;411(6837):599–603. doi: 10.1038/35079107 [published Online First: 2001/06/01]

8. Hugot JP, Laurent-Puig P, Gower-Rousseau C, et al. Mapping of a susceptibility locus for Crohn’s disease on chromosome 16. Nature 1996;379(6568):821–3. doi: 10.1038/379821a0 [published Online First: 1996/02/29]

9. Satsangi J, Parkes M, Louis E, et al. Two stage genome-wide search in inflammatory bowel disease provides evidence for susceptibility loci on chromosomes 3, 7 and 12. Nat Genet 1996;14(2):199–202. doi: 10.1038/ng1096-199 [published Online First: 1996/10/01]

10. Gettler K, Levantovsky R, Moscati A, et al. Common and Rare Variant Prediction and Penetrance of IBD in a Large, Multi-ethnic, Health System-based Biobank Cohort. Gastroenterology 2021;160(5):1546–57. doi: 10.1053/j.gastro.2020.12.034 [published Online First: 2020/12/29]

11. Piovani D, Danese S, Peyrin-Biroulet L, et al. Environmental Risk Factors for Inflammatory Bowel Diseases: An Umbrella Review of Meta-analyses. Gastroenterology 2019;157(3):647-59.e4. doi: 10.1053/j.gastro.2019.04.016 [published Online First: 2019/04/25]

12. Casey K, Lopes EW, Niccum B, et al. Alcohol consumption and risk of inflammatory bowel disease among three prospective US cohorts. Aliment Pharmacol Ther 2022;55(2):225–33. doi: 10.1111/apt.16731 [published Online First: 2021/12/10]

13. Orr WC, Fass R, Sundaram SS, et al. The effect of sleep on gastrointestinal functioning in common digestive diseases. Lancet Gastroenterol Hepatol 2020;5(6):616–24. doi: 10.1016/s2468-1253(19)30412-1 [published Online First: 2020/05/18]

14. Sudlow C, Gallacher J, Allen N, et al. UK biobank: an open access resource for identifying the causes of a wide range of complex diseases of middle and old age. PLoS medicine 2015;12(3):e1001779. doi: 10.1371/journal.pmed.1001779 [published Online First: 2015/04/01]

15. Privé F, Arbel J, Vilhjálmsson BJ. LDpred2: better, faster, stronger. Bioinformatics (Oxford, England) 2020;36(22-23):5424–31. doi: 10.1093/bioinformatics/btaa1029 [published Online First: 2020/12/17]

16. van der Sloot KWJ, Weersma RK, Alizadeh BZ, et al. Identification of Environmental Risk Factors Associated With the Development of Inflammatory Bowel Disease. Journal of Crohn’s & colitis 2020;14(12):1662–71. doi: 10.1093/ecco-jcc/jjaa114 [published Online First: 2020/06/24]

17. Satsangi J, Silverberg MS, Vermeire S, et al. The Montreal classification of inflammatory bowel disease: controversies, consensus, and implications. Gut 2006;55(6):749–53. doi: 10.1136/gut.2005.082909 [published Online First: 2006/05/16]

18. Fine JP, Gray RJ. A Proportional Hazards Model for the Subdistribution of a Competing Risk. Journal of the American Statistical Association 1999;94(446):496–509. doi: 10.1080/01621459.1999.10474144

19. Khalili H, Ananthakrishnan AN, Konijeti GG, et al. Physical activity and risk of inflammatory bowel disease: prospective study from the Nurses’ Health Study cohorts. Bmj 2013;347:f6633. doi: 10.1136/bmj.f6633 [published Online First: 2013/11/16]

20. Rahmani J, Kord-Varkaneh H, Hekmatdoost A, et al. Body mass index and risk of inflammatory bowel disease: A systematic review and dose-response meta-analysis of cohort studies of over a million participants. Obes Rev 2019;20(9):1312–20. doi: 10.1111/obr.12875 [published Online First: 2019/06/14]

21. Lo CH, Khalili H, Song M, et al. Healthy Lifestyle Is Associated With Reduced Mortality in Patients With Inflammatory Bowel Diseases. Clin Gastroenterol Hepatol 2021;19(1):87-95.e4. doi: 10.1016/j.cgh.2020.02.047 [published Online First: 2020/03/07]

22. Mahid SS, Minor KS, Soto RE, et al. Smoking and inflammatory bowel disease: a meta-analysis. Mayo Clin Proc 2006;81(11):1462–71. doi: 10.4065/81.11.1462

23. Higuchi LM, Khalili H, Chan AT, et al. A prospective study of cigarette smoking and the risk of inflammatory bowel disease in women. Am J Gastroenterol 2012;107(9):1399–406. doi: 10.1038/ajg.2012.196 [published Online First: 2012/07/11]

24. Salih A, Widbom L, Hultdin J, et al. Smoking is associated with risk for developing inflammatory bowel disease including late onset ulcerative colitis: a prospective study. Scand J Gastroenterol 2018;53(2):173–78. doi: 10.1080/00365521.2017.1418904 [published Online First: 20171221]

25. Nishikawa A, Tanaka K, Miyake Y, et al. Active and passive smoking and risk of ulcerative colitis: A case-control study in Japan. J Gastroenterol Hepatol 2021 doi: 10.1111/jgh.15745 [published Online First: 20211129]

26. To N, Gracie DJ, Ford AC. Systematic review with meta-analysis: the adverse effects of tobacco smoking on the natural history of Crohn’s disease. Aliment Pharmacol Ther 2016;43(5):549–61. doi: 10.1111/apt.13511 [published Online First: 20160107]

27. To N, Ford AC, Gracie DJ. Systematic review with meta-analysis: the effect of tobacco smoking on the natural history of ulcerative colitis. Aliment Pharmacol Ther 2016;44(2):117–26. doi: 10.1111/apt.13663 [published Online First: 20160518]

28. Aldhous MC, Drummond HE, Anderson N, et al. Smoking habit and load influence age at diagnosis and disease extent in ulcerative colitis. Am J Gastroenterol 2007;102(3):589–97. doi: 10.1111/j.1572-0241.2007.01065.x [published Online First: 2007/03/07]

29. Milajerdi A, Abbasi F, Esmaillzadeh A. A systematic review and meta-analysis of prospective studies on obesity and risk of inflammatory bowel disease. Nutr Rev 2021 doi: 10.1093/nutrit/nuab028 [published Online First: 2021/06/23]

30. Chan SSM, Chen Y, Casey K, et al. Obesity is Associated With Increased Risk of Crohn’s disease, but not Ulcerative Colitis: A Pooled Analysis of Five Prospective Cohort Studies. Clin Gastroenterol Hepatol 2021 doi: 10.1016/j.cgh.2021.06.049 [published Online First: 20210707]

31. Carreras-Torres R, Ibáñez-Sanz G, Obón-Santacana M, et al. Identifying environmental risk factors for inflammatory bowel diseases: a Mendelian randomization study. Sci Rep 2020;10(1):19273. doi: 10.1038/s41598-020-76361-2 [published Online First: 2020/11/08]

32. Wang Q, Xu KQ, Qin XR, et al. Association between physical activity and inflammatory bowel disease risk: A meta-analysis. Dig Liver Dis 2016;48(12):1425–31. doi: 10.1016/j.dld.2016.08.129 [published Online First: 2016/09/28]

33. Khalili H, Håkansson N, Chan SS, et al. Adherence to a Mediterranean diet is associated with a lower risk of later-onset Crohn’s disease: results from two large prospective cohort studies. Gut 2020;69(9):1637–44. doi: 10.1136/gutjnl-2019-319505 [published Online First: 20200103]

34. Ananthakrishnan AN, Khalili H, Konijeti GG, et al. Sleep duration affects risk for ulcerative colitis: a prospective cohort study. Clin Gastroenterol Hepatol 2014;12(11):1879–86. doi: 10.1016/j.cgh.2014.04.021 [published Online First: 2014/05/02]

35. Yadav P, Ellinghaus D, Rémy G, et al. Genetic Factors Interact With Tobacco Smoke to Modify Risk for Inflammatory Bowel Disease in Humans and Mice. Gastroenterology 2017;153(2):550–65. doi: 10.1053/j.gastro.2017.05.010 [published Online First: 2017/05/17]

36. Bridger S, Lee JC, Bjarnason I, et al. In siblings with similar genetic susceptibility for inflammatory bowel disease, smokers tend to develop Crohn’s disease and non-smokers develop ulcerative colitis. Gut 2002;51(1):21–5. doi: 10.1136/gut.51.1.21 [published Online First: 2002/06/22]

37. Ng SC, Woodrow S, Patel N, et al. Role of genetic and environmental factors in British twins with inflammatory bowel disease. Inflamm Bowel Dis 2012;18(4):725–36. doi: 10.1002/ibd.21747 [published Online First: 2011/05/11]

38. Khalili H, Chan SSM, Lochhead P, et al. The role of diet in the aetiopathogenesis of inflammatory bowel disease. Nat Rev Gastroenterol Hepatol 2018;15(9):525–35. doi: 10.1038/s41575-018-0022-9 [published Online First: 2018/05/24]

