## Supplementary for "The contribution of genetic risk and lifestyle factors in the development of adult-onset inflammatory bowel disease: a prospective cohort study"

**Supplementary Methods**

**Supplementary Table 1.** Genetic variants associated with Crohn’s disease and ulcerative colitis

**Supplementary Table 2.** Definitions of lifestyle factors

**Supplementary Table 3.** Definitions of major depressive disorder

**Supplementary Table 4.** Risk of incident Crohn's disease and ulcerative colitis according to polygenic risk score and genomic risk score

**Supplementary Table 5.** Associations between the polygenic risk score of Crohn’s disease, ulcerative colitis and lifestyle factors

**Supplementary Table 6.** Risk of incident Crohn's disease and ulcerative colitis according to alcohol consumption

**Supplementary Table 7.** Risk of incident Crohn's disease and ulcerative colitis with smoking status

**Supplementary Table 8.** Risk of incident Crohn’s disease of different disease locations with smoking status

**Supplementary Table 9.** Risk of incident ulcerative colitis of different disease locations with smoking status

**Supplementary Table 10.** Associations between smoking status and Crohn’s disease and ulcerative colitis

**Supplementary Table 11.** Associations between smoking status and Crohn’s disease and ulcerative colitis stratified by age of onset

**Supplementary Table 12**. Risk of incident Crohn's disease and ulcerative colitis by number of healthy lifestyle factors

**Supplementary Table 13**. Risk of incident Crohn's disease and ulcerative colitis by unweighted healthy lifestyle categories

**Supplementary Table 14**. Interactions of lifestyle factors and healthy lifestyle category with genetic risk

**Supplementary Table 15**. Risk of incident Crohn’s disease by genetic and lifestyle risk with unweighted lifestyle score, and unweighted and weighted lifestyle score in multivariate imputed data, and after excluding participants with incomplete data of covariables

**Supplementary Table 16**. Risk of incident ulcerative colitis by genetic and lifestyle risk with unweighted lifestyle score, and unweighted and weighted lifestyle score in multivariate imputed data, and after excluding participants with incomplete data of covariables

**Supplementary Table 17**. Risk of Incident Crohn’s disease by genetic and lifestyle risk after excluding incident Crohn’s disease within 2, 3 years after baseline, after excluding baseline colorectal cancer, and additionally adjusted for depression

**Supplementary Table 18**. Risk of Incident ulcerative colitis by genetic and lifestyle risk after excluding incident ulcerative colitis within 2, 3 years after baseline, after excluding baseline colorectal cancer, and additionally adjusted for depression

**Supplementary Table 19**. Risk of Incident Crohn’s disease by genetic and lifestyle risk stratified by covariates

**Supplementary Table 20**. Risk of Incident ulcerative colitis by genetic and lifestyle risk stratified by covariates

**Supplementary Figure 1**. Distribution of the polygenic risk score for Crohn’s disease (A) and ulcerative colitis (B)

**Supplementary Methods**

**Lifestyle factors**

Specifically, smoking status was defined as never or ever smoking, and smoking frequency (only occasionally, most or all day) of previous and current smokers was also considered in further subgroup analyses. BMI was categorized as non-obese (<30.0 kg/m^2^) and obese (≥30.0 kg/m^2^) according to World Health Organization standards for the European population. Sleep duration was defined as normal (7.0-8.0 hours/day) and abnormal sleep duration (<7 or >8 hours/day).^1^ A dietary pattern including 7 common foods was used to assess the diet quality according to the American dietary guidelines.^2 3^ Dietary intake information was collected by a food frequency questionnaire. One diet point was given if the intakes were met for a) fruits ≥3 times/day; b) vegetables ≥3 times/day; c) fish ≥2 times/week; d) whole grains ≥3 times/day; e) refined grains ≤1.5 times/day; f) processed meats ≤1 times/week; and g) unprocessed red meats ≤2.5 times/week. The diet score ranged from 0 to 7, and a diet score ≥4 indicated a high adherence to a healthy dietary pattern. Healthy alcohol drinking was defined as never to light alcohol consumption (0 to 14 g/d for women and 0 to 28 g/d for men) with the maximum limit reflecting the American dietary guidelines. Regular physical activity was defined by at least ≥150 minutes moderate activity per week or ≥75 minutes vigorous activity per week (or an equivalent combination) or having moderate physical activity at least 5 days a week or vigorous activity once a week, as recommended by the American Heart Association.^4^

**Sensitivity analysis**

The associations were further examined with several sensitivity analyses, including: 1) an analysis using the genetic risk quintiles instead of categories; 2) an analysis using the number of healthy lifestyle factors instead of categories; 3) an analysis using an unweighted lifestyle score; 4) an analysis excluding baseline colorectal cancer patients; 5) an analysis excluding participants with incomplete covariate data; and 6) with further adjustment for baseline major depressive disorder.

**References**

1. Ananthakrishnan AN, Khalili H, Konijeti GG, et al. Sleep duration affects risk for ulcerative colitis: a prospective cohort study. *Clin Gastroenterol Hepatol* 2014;12(11):1879-86. doi: 10.1016/j.cgh.2014.04.021 [published Online First: 2014/05/02]

2. Mozaffarian D, Appel LJ, Van Horn L. Components of a cardioprotective diet: new insights. *Circulation* 2011;123(24):2870-91. doi: 10.1161/circulationaha.110.968735 [published Online First: 2011/06/22]

3. US Department of Health and Human Services. 2015-2020 Dietary guidelines for Americans. 8th edition. December 2015.

4. Lloyd-Jones DM, Hong Y, Labarthe D, et al. Defining and setting national goals for cardiovascular health promotion and disease reduction: the American Heart Association's strategic Impact Goal through 2020 and beyond. *Circulation* 2010;121(4):586-613. doi: 10.1161/circulationaha.109.192703 [published Online First: 2010/01/22]

**Supplementary Table 1.** Genetic variants associated with Crohn’s disease and ulcerative colitis

| **Outcome** | **SNP** | **Chr** | **Pos_hg19** | **EA** | **NEA** | **Beta** | **SE** | **Pval** |
| --- | --- | --- | --- | --- | --- | --- | --- | --- |
| CD | rs7517847 | 1 | 67681669 | C | A | -0.342 | 0.017 | 2.29×10^-46 |
| CD | rs7517810 | 1 | 172853460 | A | G | 0.131 | 0.012 | 1.11×10^-14 |
| CD | rs3024505 | 1 | 206939904 | A | G | 0.166 | 0.014 | 3.91×10^-09 |
| CD | rs10798069 | 1 | 186875459 | A | C | -0.073 | 0.011 | 4.25×10^-09 |
| CD | rs7555082 | 1 | 198598663 | A | G | 0.122 | 0.018 | 1.47×10^-10 |
| CD | rs10495903 | 2 | 43806918 | A | G | 0.122 | 0.015 | 3.3×10^-08 |
| CD | rs6708413 | 2 | 103063369 | G | A | 0.113 | 0.013 | 1.54×10^-10 |
| CD | rs6716753 | 2 | 231097129 | G | A | 0.131 | 0.013 | 1.87×10^-08 |
| CD | rs12994997 | 2 | 234173503 | G | A | -0.223 | 0.015 | 2.06×10^-37 |
| CD | rs11681525 | 2 | 145492382 | C | G | -0.151 | 0.024 | 4.08×10^-11 |
| CD | rs35320439 | 2 | 242737341 | G | A | 0.086 | 0.009 | 9.89×10^-10 |
| CD | rs3197999 | 3 | 49721532 | A | G | 0.157 | 0.011 | 9.13×10^-13 |
| CD | rs11742570 | 5 | 40410584 | A | G | -0.248 | 0.016 | 4.47×10^-34 |
| CD | rs1363907 | 5 | 96252803 | A | G | 0.104 | 0.012 | 1.46×10^-11 |
| CD | rs11743851 | 5 | 130613600 | G | A | 0.140 | 0.011 | 8.69×10^-12 |
| CD | rs11741861 | 5 | 150277909 | G | A | 0.285 | 0.015 | 4.6×10^-16 |
| CD | rs6556412 | 5 | 158787385 | A | G | 0.157 | 0.011 | 8.03×10^-15 |
| CD | rs6908425 | 6 | 20728731 | A | G | -0.105 | 0.017 | 2.42×10^-08 |
| CD | rs7746082 | 6 | 106435269 | C | G | 0.131 | 0.012 | 1.77×10^-08 |
| CD | rs1819333 | 6 | 167373547 | C | A | -0.117 | 0.014 | 9.3×10^-12 |
| CD | rs7773324 | 6 | 382559 | G | A | -0.083 | 0.011 | 1.06×10^-09 |
| CD | rs13204048 | 6 | 3420406 | G | A | -0.073 | 0.011 | 2.89×10^-08 |
| CD | rs7758080 | 6 | 149577079 | G | A | 0.077 | 0.009 | 7.27×10^-09 |
| CD | rs1456896 | 7 | 50304461 | G | A | -0.094 | 0.014 | 2.9×10^-08 |
| CD | rs921720 | 8 | 126534671 | A | G | -0.117 | 0.014 | 6.4×10^-12 |
| CD | rs4246905 | 9 | 117553249 | A | G | -0.139 | 0.016 | 1.28×10^-14 |
| CD | rs10781499 | 9 | 139266405 | A | G | 0.166 | 0.010 | 1.03×10^-19 |
| CD | rs11010067 | 10 | 35295431 | G | C | 0.131 | 0.011 | 2.33×10^-09 |
| CD | rs10761659 | 10 | 64445564 | A | G | -0.186 | 0.015 | 3.42×10^-19 |
| CD | rs4409764 | 10 | 101284237 | A | C | 0.174 | 0.010 | 8.48×10^-19 |
| CD | rs2155219 | 11 | 76299194 | A | C | 0.174 | 0.010 | 6.51×10^-13 |
| CD | rs12422544 | 12 | 40528432 | G | A | 0.378 | 0.025 | 3.29×10^-12 |
| CD | rs7954567 | 12 | 6491125 | A | G | 0.086 | 0.009 | 1.3×10^-09 |
| CD | rs3764147 | 13 | 44457925 | G | A | 0.140 | 0.012 | 7.31×10^-09 |
| CD | rs9525625 | 13 | 43018030 | A | G | 0.077 | 0.009 | 1.41×10^-09 |
| CD | rs17293632 | 15 | 67442596 | A | G | 0.131 | 0.012 | 1.08×10^-12 |
| CD | rs26528 | 16 | 28517709 | G | A | 0.122 | 0.011 | 1.06×10^-08 |
| CD | rs3091315 | 17 | 32593665 | G | A | -0.139 | 0.016 | 9.52×10^-12 |
| CD | rs12946510 | 17 | 37912377 | A | G | 0.122 | 0.011 | 4.3×10^-08 |
| CD | rs3853824 | 17 | 54880993 | A | G | -0.083 | 0.011 | 1.17×10^-10 |
| CD | rs1893217 | 18 | 12809340 | G | A | 0.166 | 0.014 | 1.92×10^-12 |
| CD | rs7236492 | 18 | 77220616 | A | G | -0.094 | 0.022 | 9.09×10^-09 |
| CD | rs2024092 | 19 | 1124031 | A | G | 0.148 | 0.012 | 2.26×10^-11 |
| CD | rs11879191 | 19 | 10512911 | A | G | -0.139 | 0.019 | 1.66×10^-08 |
| CD | rs516246 | 19 | 49206172 | A | G | 0.113 | 0.011 | 1.21×10^-08 |
| CD | rs6062504 | 20 | 62348907 | A | G | -0.105 | 0.015 | 3.28×10^-10 |
| CD | rs2823286 | 21 | 16817938 | A | G | -0.139 | 0.015 | 1.24×10^-09 |
| CD | rs7282490 | 21 | 45615741 | G | A | 0.122 | 0.011 | 3.81×10^-13 |
| CD | rs2256609 | 22 | 21925017 | G | A | 0.104 | 0.014 | 8.02×10^-09 |
| CD | rs2413583 | 22 | 39659773 | A | G | -0.211 | 0.021 | 3.65×10^-10 |
| CD | rs727563 | 22 | 41867377 | G | A | 0.095 | 0.009 | 1.88×10^-10 |
| UC | rs6667605 | 1 | 2502780 | A | G | -0.083 | 0.014 | 1.4×10^-08 |
| UC | rs3806308 | 1 | 20142866 | A | G | -0.174 | 0.016 | 9.81×10^-15 |
| UC | rs6426833 | 1 | 20171860 | G | A | -0.236 | 0.017 | 4.86×10^-31 |
| UC | rs12568930 | 1 | 22702231 | G | A | -0.128 | 0.021 | 6.24×10^-11 |
| UC | rs7517847 | 1 | 67681669 | C | A | -0.151 | 0.015 | 6.03×10^-11 |
| UC | rs1801274 | 1 | 161479745 | G | A | -0.174 | 0.016 | 3.78×10^-17 |
| UC | rs7554511 | 1 | 200877562 | A | C | -0.163 | 0.017 | 1.05×10^-11 |
| UC | rs3024505 | 1 | 206939904 | A | G | 0.223 | 0.013 | 2.97×10^-17 |
| UC | rs7608910 | 2 | 61204856 | G | A | 0.131 | 0.012 | 1.81×10^-12 |
| UC | rs3749171 | 2 | 241569692 | A | G | 0.140 | 0.014 | 2.33×10^-10 |
| UC | rs9868809 | 3 | 48681053 | A | G | 0.148 | 0.018 | 6.01×10^-09 |
| UC | rs113010081 | 3 | 46457412 | G | A | 0.131 | 0.018 | 9.02×10^-10 |
| UC | rs2189234 | 4 | 106075498 | A | C | 0.077 | 0.009 | 1.95×10^-10 |
| UC | rs254560 | 5 | 134443606 | A | G | 0.077 | 0.012 | 1.6×10^-08 |
| UC | rs56167332 | 5 | 158827769 | A | C | 0.140 | 0.011 | 5.3×10^-11 |
| UC | rs6920220 | 6 | 138006504 | A | G | 0.148 | 0.013 | 5.22×10^-11 |
| UC | rs7805114 | 7 | 107450033 | C | A | -0.117 | 0.015 | 5.86×10^-09 |
| UC | rs1077773 | 7 | 17442679 | G | A | -0.073 | 0.011 | 5.96×10^-09 |
| UC | rs4246905 | 9 | 117553249 | A | G | -0.117 | 0.016 | 1.97×10^-08 |
| UC | rs10781499 | 9 | 139266405 | A | G | 0.131 | 0.012 | 7.69×10^-13 |
| UC | rs4409764 | 10 | 101284237 | A | C | 0.157 | 0.011 | 8.39×10^-15 |
| UC | rs2155219 | 11 | 76299194 | A | C | 0.122 | 0.012 | 1.48×10^-09 |
| UC | rs483905 | 11 | 96023427 | A | G | 0.086 | 0.013 | 1.57×10^-08 |
| UC | rs561722 | 11 | 114386830 | A | G | -0.128 | 0.016 | 5.21×10^-09 |
| UC | rs7134472 | 12 | 68499986 | A | G | 0.157 | 0.011 | 4.77×10^-18 |
| UC | rs17085007 | 13 | 27531267 | G | A | 0.131 | 0.014 | 1.18×10^-08 |
| UC | rs12946510 | 17 | 37912377 | A | G | 0.131 | 0.012 | 4.95×10^-10 |
| UC | rs17736589 | 17 | 76737118 | G | A | 0.086 | 0.019 | 4.34×10^-08 |
| UC | rs2836878 | 21 | 40465534 | A | G | -0.223 | 0.019 | 2.05×10^-20 |
| UC | rs7282490 | 21 | 45615741 | G | A | 0.104 | 0.012 | 7.08×10^-11 |

CD, Crohn's disease; Chr, chromosome; EA, effect allele; NEA, non-effect allele; SNP; single nucleotide polymorphism; UC, ulcerative colitis.

**l**

**Supplementary Table 2.** Definitions of lifestyle factors

| **Lifestyle factors** | **Definitions** | **UK Biobank field code** |
| --- | --- | --- |
| **Smoking** | 1-Ever smoking (both previous and current)  UK Biobank Touchscreen questionnaire at baseline divided the smoking status into three categories: never, previous and current | 20116 |
| **BMI** | 1-BMI ≥18.5 and <25.0  UK Biobank measured body composition manually at baseline, and BMI was constructed from height and weight measured (weight/height-square, Kg/m2) | 21001 |
| **Sleep** | 1-Sleep duration ≥7 and ≤8  UK Biobank Touchscreen question 'About how many hours sleep do you get in every 24 hours? (Please include naps)' | 1160 |
| **Diet** | 1-Consuming 4-7 adequate amounts of the recommended food groups  UK Biobank Food Frequency Questionnaire asked the frequency of intake of a range of common food and drink items.  7 recommended food groups and adequate amounts: 1. Fruits: ≥ 3 times/day 2. Vegetables: ≥ 3 times/day 3. Fish: ≥2 times/week 4. Whole grains: ≥ 3 times/day 5. Refined grains: ≤1.5 times/day 6. Processed meats: ≤ 1 times/week 7. Unprocessed red meats: ≤ 2.5 times/week | 1309, 1319, 1289, 1299, 1329, 1339, 1349, 1369, 1379, 1389, 1438, 1448, 1458, 1468 |
| **Physical activity** | 1-regular physical activity  UK Biobank Touchscreen questionnaire on the reported type and duration of physical activity (including walking, DIY, moderate and vigorous physical activity, strenuous sports, etc).  One of the following is considered as regular physical activity:  1.≥150 minutes moderate activity per week  2.≥ 75 minutes vigorous activity per week  3. Equivalent combination  4. Moderate physical activity at least 5 days a week or vigorous activity once a week | 884, 894, 904, 914 |
| **Alcohol consumption** | 1-Moderate consumption (women: >0 and ≤14g/day; men: >0 and ≤28g/day)  UK Biobank Touchscreen questionnaire on whether the participant reported drinking alcohol, frequency of intake, beverage type, whether they usually drink with meals.  According to the US Dietary Guidelines for Americans 2015-2020, up to 1 drink/day for women and up to 2 drinks/day for men. To calculate drink-equivalents as per guidelines, multiply the volume in ounces by the alcohol content in percent and divide by 0.6 ounces of alcohol per drink-equivalent; then convert to grams: 1 drink-equivalent described as containing 14g of pure alcohol.  125ml wine=0.85 drink-equivalents,  4% ABV pint beer = 1.28 drink-equivalents,  25ml spirits=0.57 drink-equivalents,  50ml fortified wine= 0.56 drink-equivalents | 1558, 1568, 1578, 1588, 1598, 1608, 5364, 4407, 4418, 4429, 4440, 4451, 4462 |

**Supplementary Table 3.** Definitions of major depressive disorder

| **ACE touchscreen question** | **Depression symptom (with score >= 3):** | **UKB Code** |
| --- | --- | --- |
| Over the past two weeks, how often have you felt down, depressed or hopeless? | 0 (not at all); 1 (several days); 2 (more than half the days); 3 (nearly every day) | 2050 |
| Over the past two weeks, how often have you had little interest or pleasure in doing things? | 0 (not at all); 1 (several days); 2 (more than half the days); 3 (nearly every day) | 2060 |

**Supplementary Table 4.** Risk of incident Crohn's disease and ulcerative colitis according to polygenic risk score and genomic risk score

|  | **Crohn's disease** | | | | |  | **Ulcerative colitis** | | | | |
| --- | --- | --- | --- | --- | --- | --- | --- | --- | --- | --- | --- |
| **Category** | **Events/**  **Person-years** | **Model 1 ^a^** |  | **Model 2 ^b^** |  |  | **Events/Person-years** | **Model 1 ^a^** |  | **Model 2 ^b^** |  |
|  |  | **HR (95% CI)** | ***P* value** | **HR (95% CI)** | ***P* value** |  |  | **HR (95% CI)** | ***P* value** | **HR (95% CI)** | ***P* value** |
| **Genetic risk (used genomic risk score)** | | | | | | | | | | | |
| Low | 115/1,006,604 | 1 (Ref) |  | 1 (Ref) |  |  | 230/1,008,599 | 1 (Ref) |  | 1 (Ref) |  |
| Intermediate | 394/3,022,021 | 1.13 (0.92, 1.40) | 0.235 | 1.14 (0.92, 1.40) | 0.229 |  | 908/3,025,318 | 1.32 (1.14, 1.52) | <0.001 | 1.92 (1.64, 2.25) | <0.001 |
| High | 198/1,007,856 | 1.70 (1.35, 2.14) | <0.001 | 1.70 (1.35, 2.14) | <0.001 |  | 438/1,008,420 | 1.92 (1.63, 2.25) | <0.001 | 1.92 (1.64, 2.25) | <0.001 |
| *p* value for trend ^c^ |  | <0.001 |  | <0.001 |  |  |  | <0.001 |  | <0.001 |  |
| **Genomic risk score (quintiles)** | | | | | | | | | | | |
| 1 (the lowest) | 115/1,006,604 | 1 (Ref) |  | 1 (Ref) |  |  | 230/1,008,599 | 1 (Ref) |  | 1 (Ref) |  |
| 2 | 109/1,007,185 | 0.94 (0.73, 1.23) | 0.666 | 0.95 (0.73, 1.23) | 0.672 |  | 251/1,008,132 | 1.09 (0.91, 1.30) | 0.344 | 1.09 (0.91, 1.30) | 0.342 |
| 3 | 136/1,008,247 | 1.17 (0.92, 1.51) | 0.205 | 1.18 (0.92, 1.51) | 0.197 |  | 299/1,008,432 | 1.30 (1.10, 1.54) | 0.003 | 1.30 (1.10, 1.54) | 0.003 |
| 4 | 149/1,006,589 | 1.29 (1.01, 1.64) | 0.044 | 1.29 (1.01, 1.64) | 0.042 |  | 358/1,008,754 | 1.57 (1.33, 1.85) | <0.001 | 1.57 (1.33, 1.85) | <0.001 |
| 5 (the highest) | 198/1,007,856 | 1.70 (1.35, 2.14) | <0.001 | 1.70 (1.35, 2.15) | <0.001 |  | 438/1,008,420 | 1.92 (1.63, 2.25) | <0.001 | 1.92 (1.64, 2.25) | <0.001 |
| *p* value for trend ^c^ |  | <0.001 |  | <0.001 |  |  |  | <0.001 |  | <0.001 |  |
| **Polygenic risk score (quintiles)** | | | | | | | | | | | |
| 1 (the lowest) | 92/1,007,314 | 1 (Ref) |  | 1 (Ref) |  |  | 212/1,009,486 | 1 (Ref) |  | 1 (Ref) |  |
| 2 | 115/1,007,410 | 1.25 (0.95, 1.64) | 0.113 | 1.25 (0.95, 1.64) | 0.114 |  | 246/1,008,916 | 1.16 (0.97, 1.39) | 0.112 | 1.16 (0.97, 1.40) | 0.108 |
| 3 | 135/1,008,087 | 1.46 (1.12, 1.90) | 0.005 | 1.46 (1.12, 1.90) | 0.005 |  | 322/1,008,795 | 1.52 (1.28, 1.81) | <0.001 | 1.52 (1.28, 1.81) | <0.001 |
| 4 | 158/1,007,491 | 1.71 (1.32, 2.21) | <0.001 | 1.71 (1.32, 2.21) | <0.001 |  | 342/1,007,759 | 1.62 (1.36, 1.92) | <0.001 | 1.62 (1.36, 1.92) | <0.001 |
| 5 (the highest) | 207/1,006,180 | 2.24 (1.75, 2.86) | <0.001 | 2.24 (1.75, 2.86) | <0.001 |  | 454/1,007,381 | 2.15 (1.83, 2.53) | <0.001 | 2.15 (1.82, 2.53) | <0.001 |
| *p* value for trend ^c^ |  |  | <0.001 |  | <0.001 |  |  |  | <0.001 |  | <0.001 |

CI indicates confidence interval; HR, hazard ratio.

^a^ Adjusted for age, age-square, sex, TDI, education, CCI, and first 20 principal components of ancestry.

^b^ Adjusted for Model 1 and weighted lifestyle categories

^c^ The trend test used the median value of each group instead of the original group.

**Supplementary Table 5.** Associations between the polygenic risk score of Crohn’s disease, ulcerative colitis and lifestyle factors

| **Lifestyle Factors** | **Polygenic risk score of CD** | | **Polygenic risk score of UC** | |
| --- | --- | --- | --- | --- |
|  | **OR (95% CI)** | ***P* value** | **OR (95% CI)** | ***P* value** |
| Never smoking | 1.01 (1.01, 1.02) | 0.002 | 1.00 (0.99, 1.01) | 0.910 |
| No obesity | 1.00 (0.99, 1.02) | 0.442 | 1.00 (0.99, 1.02) | 0.724 |
| Adequate sleep duration (7-8h) | 1.00 (0.99, 1.01) | 0.670 | 0.99 (0.98, 1.01) | 0.350 |
| Healthy diet | 0.99 (0.98, 1.00) | 0.137 | 0.99 (0.98, 1.01) | 0.324 |
| Regular physical activity | 0.99 (0.98, 1.00) | 0.212 | 1.00 (0.99, 1.02) | 0.536 |

CD, Crohn's disease; UC, ulcerative colitis; CI indicates confidence interval; OR, odds ratio.

Adjusted for age, age-square, sex, TDI, education, CCI, and first 20 principal components of ancestry, and other healthy lifestyle factors.

**Supplementary Table 6.** Risk of incident Crohn's disease and ulcerative colitis according to alcohol consumption

|  | **Crohn's disease (total n=367,703, cases n=579)** | | | | |  | **Ulcerative colitis (total n=368,474, cases n=1350)** | | | | |
| --- | --- | --- | --- | --- | --- | --- | --- | --- | --- | --- | --- |
| **Category** | **Events/**  **Person-years** | **Model 1 ^a^** |  | **Model 2 ^b^** |  |  | **Events/Person-years** | **Model 1 ^a^** |  | **Model 2 ^b^** |  |
|  |  | **HR (95% CI)** | ***P* value** | **HR (95% CI)** | ***P* value** |  |  | **HR (95% CI)** | ***P* value** | **HR (95% CI)** | ***P* value** |
| **Alcohol consumption ^c^** | |  |  |  |  |  |  |  |  |  |  |
| Moderate | 325/2,496,909 | 1 (Ref) |  | 1 (Ref) |  |  | 746/2,499,793 | 1 (Ref) |  | 1 (Ref) |  |
| Inappropriate | 254/1,791,212 | 0.99 (0.84, 1.18) | 0.946 | 0.99 (0.84, 1.18) | 0.948 |  | 604/1,793,548 | 1.03 (0.92, 1.15) | 0.632 | 1.03 (0.92, 1.15) | 0.632 |

CI indicates confidence interval; HR, hazard ratio.

^a^ Adjusted for age, age-square, sex, TDI, education, CCI, and the first 20 principal components of ancestry; and other lifestyle factors in the analysis of alcohol consumption.

^b^ Adjusted for Model 1 and genetic risk category.

**Supplementary Table 7.** Risk of incident Crohn's disease and ulcerative colitis with smoking status

| **Lifestyle Factors** | **Incident CD** | | |  | **Incident UC** | | |
| --- | --- | --- | --- | --- | --- | --- | --- |
|  | **Events/Person-years** | **HR (95% CI)** | ***P* value** |  | **Events/Person-years** | **HR (95% CI)** | ***P* value** |
| Never smoking | 324/2,760,887 | 1 (Ref) |  |  | 634/2,763,009 | 1 (Ref) |  |
| Previous smoking |  |  |  |  |  |  |  |
| Only occasionally | 60/592,582 | 0.85 (0.65, 1.12) | 0.256 |  | 166/593,224 | 1.18 (1.00, 1.41) | 0.053 |
| Most or all days | 212/1,184,480 | 1.40 (1.18, 1.68) | <0.001 |  | 559/1,186,764 | 1.81 (1.61, 2.03) | <0.001 |
| Current smoking |  |  |  |  |  |  |  |
| Only occasionally | 24/132,808 | 1.48 (0.98, 2.25) | 0.063 |  | 46/132,967 | 1.40 (1.04, 1.89) | 0.029 |
| Most or all days | 87/365,724 | 1.74 (1.36, 2.23) | <0.001 |  | 171/366,372 | 1.71 (1.44, 2.04) | <0.001 |

CI indicates confidence interval; HR, hazard ratio; PAR, population attributable risk.

Adjusted for age, age-square, sex, TDI, education, CCI, polygenic risk score of CD or UC, and first 20 principal components of ancestry, and other lifestyle factors.

**Supplementary Table 8.** Risk of incident Crohn’s disease of different disease locations with smoking status

| **Smoking status** | **L1 (Small bowel disease or terminal ileitis)** | | |  | **L2 (Colon)** | | |
| --- | --- | --- | --- | --- | --- | --- | --- |
|  | **Events/Person-years** | **HR (95% CI)** | ***P* value** |  | **Events/Person-years** | **HR (95% CI)** | ***P* value** |
| Never smoking | 49/2,759,089 | 1 (Ref) |  |  | 40/2,759,017 | 1 (Ref) |  |
| Previous smoking |  |  |  |  |  |  |  |
| Only occasionally | 8/592,230 | 0.78 (0.37, 1.64) | 0.511 |  | 6/592,198 | 0.72 (0.30, 1.70) | 0.452 |
| Most or all days | 50/1,183,482 | 2.34 (1.56, 3.51) | <0.001 |  | 23/1,183,270 | 1.29 (0.76, 2.18) | 0.345 |
| Current smoking |  |  |  |  |  |  |  |
| Only occasionally | 4/132,691 | 1.62 (0.58, 4.50) | 0.358 |  | 3/132,664 | 1.56 (0.48, 5.08) | 0.459 |
| Most or all days | 23/365,316 | 3.18 (1.90, 5.35) | <0.001 |  | 11/365,252 | 1.92 (0.96, 3.85) | 0.066 |
| **Smoking status** | **L3/LX (Ileocecal or location not defined)** | | |  |  | | |
|  | **Events/Person-years** | **HR (95% CI)** | ***P* value** |  |  |  |  |
| Never smoking | 235/2,760,293 | 1 (Ref) |  |  |  |  |  |
| Previous smoking |  |  |  |  |  |  |  |
| Only occasionally | 46/592,490 | 0.89 (0.65, 1.22) | 0.458 |  |  |  |  |
| Most or all days | 139/1,183,975 | 1.24 (1.00, 1.54) | 0.051 |  |  |  |  |
| Current smoking |  |  |  |  |  |  |  |
| Only occasionally | 17/132,771 | 1.45 (0.88, 2.37) | 0.145 |  |  |  |  |
| Most or all days | 53/365,532 | 1.43 (1.05, 1.95) | 0.024 |  |  |  |  |

CD, Crohn’s disease; CI indicates confidence interval; HR, hazard ratio.

Adjusted for age, age-square, sex, TDI, education, CCI, polygenic risk score of CD, and first 20 principal components of ancestry, and other lifestyle factors.

**Supplementary Table 9.** Risk of incident ulcerative colitis of different disease locations with smoking status

| **Smoking status** | **E1 (Ulcerative proctitis)** | | |  | **E2 (Left-sided UC)** | | |
| --- | --- | --- | --- | --- | --- | --- | --- |
|  | **Events/Person-years** | **HR (95% CI)** | ***P* value** |  | **Events/Person-years** | **HR (95% CI)** | ***P* value** |
| Never smoking | 72/2,759,230 | 1 (Ref) |  |  | 59/2,759,189 | 1 (Ref) |  |
| Previous smoking |  |  |  |  |  |  |  |
| Only occasionally | 17/592,288 | 1.09 (0.64, 1.85) | 0.752 |  | 10/592,228 | 0.75 (0.39, 1.47) | 0.409 |
| Most or all days | 46/1,183,428 | 1.46 (1.00, 2.13) | 0.051 |  | 57/1,183,518 | 1.90 (1.31, 2.76) | 0.001 |
| Current smoking |  |  |  |  |  |  |  |
| Only occasionally | 5/132,688 | 1.29 (0.52, 3.22) | 0.578 |  | 4/132,692 | 1.24 (0.45, 3.43) | 0.678 |
| Most or all days | 9/365,258 | 0.84 (0.41, 1.71) | 0.631 |  | 18/365,333 | 1.83 (1.06, 3.17) | 0.031 |
| **Smoking status** | **E3 (Extensive UC)** | | |  | **EX (Extent not defined)** | | |
|  | **Events/Person-years** | **HR (95% CI)** | ***P* value** |  | **Events/Person-years** | **HR (95% CI)** | ***P* value** |
| Never smoking | 36/2,759,047 | 1 (Ref) |  |  | 467/2,761,812 | 1 (Ref) |  |
| Previous smoking |  |  |  |  |  |  |  |
| Only occasionally | 6/592,219 | 0.74 (0.31, 1.76) | 0.500 |  | 133/592,993 | 1.29 (1.06, 1.56) | 0.010 |
| Most or all days | 36/1,183,413 | 2.05 (1.27, 3.29) | 0.003 |  | 420/1,185,776 | 1.83 (1.60, 2.09) | <0.001 |
| Current smoking |  |  |  |  |  |  |  |
| Only occasionally | 2/132,673 | 1.15 (0.28, 4.82) | 0.844 |  | 35/132,891 | 1.45 (1.03, 2.05) | 0.035 |
| Most or all days | 13/365,288 | 2.65 (1.37, 5.13) | 0.004 |  | 131/366,056 | 1.76 (1.44, 2.15) | <0.001 |

UC, ulcerative colitis; CI indicates confidence interval; HR, hazard ratio.

Adjusted for age, age-square, sex, TDI, education, CCI, polygenic risk score of UC, and first 20 principal components of ancestry, and other lifestyle factors.

**Supplementary Table 10.** Associations between smoking status and Crohn’s disease and ulcerative colitis

| **Smoking status** | **CD (cases, n=2343)** | | | | **UC (cases, n=5130)** | | | |
| --- | --- | --- | --- | --- | --- | --- | --- | --- |
|  | **Crude model** | | **Fully adjusted** | | **Crude model** | | **Fully adjusted** | |
|  | **OR (95%CI)** | ***P* value** | **OR (95%CI)** | ***P* value** | **OR (95%CI)** | ***P* value** | **OR (95%CI)** | ***P* value** |
| Never smoking | 1 (Ref) |  | 1 (Ref) |  | 1 (Ref) |  | 1 (Ref) |  |
| Ever smoking | 1.47 (1.35, 1.59) | <0.001 | 1.41 (1.29, 1.53) | <0.001 | 1.46 (1.38, 1.54) | <0.001 | 1.36 (1.28, 1.44) | <0.001 |
| Previous smoking | 1.39 (1.27, 1.52) | <0.001 | 1.38 (1.26, 1.50) | <0.001 | 1.57 (1.48, 1.66) | <0.001 | 1.47 (1.39, 1.56) | <0.001 |
| Current smoking | 1.74 (1.54, 1.97) | <0.001 | 1.51 (1.33, 1.71) | <0.001 | 1.08 (0.97, 1.19) | 0.156 | 0.96 (0.86, 1.06) | 0.418 |

CD, Crohn's disease; UC, ulcerative colitis; CI indicates confidence interval; OR, odds ratio.

Adjusted for age, age-square, sex, TDI, education, polygenic risk score of CD or UC, and first 20 principal components of ancestry, and other lifestyle factors.

**Supplementary Table 11.** Associations between smoking status and Crohn’s disease and ulcerative colitis stratified by age of onset

| **Subgroup** | **Crohn’s disease** | | | | **Ulcerative colitis** | | | |
| --- | --- | --- | --- | --- | --- | --- | --- | --- |
|  | **Crude model** | | **Fully adjusted** | | **Crude model** | | **Fully adjusted** | |
|  | **OR (95%CI)** | ***P* value** | **OR (95%CI)** | ***P* value** | **OR (95%CI)** | ***P* value** | **OR (95%CI)** | ***P* value** |
| **Aged <= 20 years old (Number of cases, CD n=155; UC n = 216)** | | | | | | | | |
| Never smoking | 1 (Ref) |  | 1 (Ref) |  | 1 (Ref) |  | 1 (Ref) |  |
| Ever smoking | 1.27 (0.93, 1.74) | 0.135 | 1.27 (0.92, 1.75) | 0.149 | 0.70 (0.53, 0.92) | 0.012 | 0.74 (0.56, 0.97) | 0.032 |
| Previous smoking | 1.33 (0.95, 1.85) | 0.095 | 1.39 (0.99, 1.95) | 0.054 | 0.65 (0.48, 0.89) | 0.007 | 0.69 (0.50, 0.94) | 0.018 |
| Current smoking | 1.08 (0.62, 1.87) | 0.794 | 0.88 (0.50, 1.56) | 0.669 | 0.87 (0.56, 1.37) | 0.548 | 0.92 (0.58, 1.46) | 0.715 |
| **Aged 20-40 years old (Number of cases, CD n=739; UC n = 1452)** | | | | | | | | |
| Never smoking | 1 (Ref) |  | 1 (Ref) |  | 1 (Ref) |  | 1 (Ref) |  |
| Ever smoking | 1.31 (1.13, 1.51) | <0.001 | 1.31 (1.13, 1.52) | <0.001 | 0.99 (0.89, 1.10) | 0.860 | 0.99 (0.89, 1.10) | 0.818 |
| Previous smoking | 1.19 (1.02, 1.40) | 0.029 | 1.24 (1.06, 1.45) | 0.009 | 1.09 (0.98, 1.22) | 0.100 | 1.10 (0.99, 1.23) | 0.079 |
| Current smoking | 1.73 (1.40, 2.14) | <0.001 | 1.57 (1.26, 1.95) | <0.001 | 0.62 (0.50, 0.78) | <0.001 | 0.58 (0.47, 0.72) | <0.001 |
| **Aged 40-60 years old (Number of cases, CD n=846; UC n = 2034)** | | | | | | | | |
| Never smoking | 1 (Ref) |  | 1 (Ref) |  | 1 (Ref) |  | 1 (Ref) |  |
| Ever smoking | 1.60 (1.40, 1.84) | <0.001 | 1.51 (1.31, 1.73) | <0.001 | 1.68 (1.54, 1.84) | <0.001 | 1.57 (1.44, 1.72) | <0.001 |
| Previous smoking | 1.44 (1.24, 1.67) | <0.001 | 1.41 (1.22, 1.64) | <0.001 | 1.80 (1.64, 1.98) | <0.001 | 1.71 (1.56, 1.88) | <0.001 |
| Current smoking | 2.16 (1.78, 2.62) | <0.001 | 1.83 (1.50, 2.24) | <0.001 | 1.25 (1.07, 1.46) | 0.005 | 1.07 (0.91, 1.26) | 0.390 |
| **Aged > 60 years old (Number of cases, CD n=603, UC n = 1428)** | | | | | | | | |
| Never smoking | 1 (Ref) |  | 1 (Ref) |  | 1 (Ref) |  | 1 (Ref) |  |
| Ever smoking | 1.55 (1.32, 1.82) | <0.001 | 1.45 (1.23, 1.70) | <0.001 | 1.99 (1.79, 2.22) | <0.001 | 1.83 (1.64, 2.04) | <0.001 |
| Previous smoking | 1.61 (1.36, 1.90) | <0.001 | 1.53 (1.29, 1.81) | <0.001 | 2.12 (1.90, 2.37) | <0.001 | 1.97 (1.76, 2.21) | <0.001 |
| Current smoking | 1.36 (1.04, 1.78) | 0.027 | 1.16 (0.88, 1.54) | 0.289 | 1.52 (1.27, 1.82) | <0.001 | 1.29 (1.07, 1.56) | 0.007 |

CD, Crohn's disease; UC, ulcerative colitis.

Fully adjusted for sex, TDI, education, polygenic risk score of CD or UC, and first 20 principal components of ancestry, and other lifestyle factors.

**Supplementary Table 12**. Risk of incident Crohn's disease and ulcerative colitis by number of healthy lifestyle factors

| **Number of healthy lifestyle factors** | **Events/Person-years** | **Model 1 ^a^** | | **Model 2 ^b^** | |
| --- | --- | --- | --- | --- | --- |
|  |  | **HR (95% CI)** | ***P* value** | **HR (95% CI)** | ***P* value** |
| Crohn's disease |  |  |  |  |  |
| 5 | 94/967,897 | 1 (Ref) |  | 1 (Ref) |  |
| 4 | 192/1,728,927 | 1.11 (0.87, 1.42) | 0.402 | 1.11 (0.87, 1.42) | 0.402 |
| 3 | 209/1,395,891 | 1.45 (1.14, 1.86) | 0.003 | 1.46 (1.14, 1.86) | 0.003 |
| 2 | 144/693,449 | 1.95 (1.50, 2.54) | <0.001 | 1.95 (1.50, 2.55) | <0.001 |
| 1 | 57/217,805 | 2.37 (1.69, 3.32) | <0.001 | 2.37 (1.69, 3.33) | <0.001 |
| 0 | 11/32,512 | 3.00 (1.60, 5.64) | 0.001 | 3.02 (1.61, 5.67) | 0.001 |
| p value for trend c |  |  | <0.001 |  | <0.001 |
| Ulcerative colitis |  |  |  |  |  |
| 5 | 198/968,658 |  |  |  |  |
| 4 | 443/1,730,576 | 1.18 (1.00, 1.40) | 0.051 | 1.18 (1.00, 1.40) | 0.052 |
| 3 | 482/1,397,726 | 1.50 (1.27, 1.78) | <0.001 | 1.50 (1.27, 1.78) | <0.001 |
| 2 | 321/694,616 | 1.90 (1.59, 2.28) | <0.001 | 1.90 (1.59, 2.28) | <0.001 |
| 1 | 117/218,201 | 2.11 (1.67, 2.66) | <0.001 | 2.10 (1.66, 2.65) | <0.001 |
| 0 | 15/32,559 | 1.75 (1.03, 2.96) | 0.039 | 1.74 (1.03, 2.95) | 0.040 |
| p value for trend c |  |  | <0.001 |  | <0.001 |

CI indicates confidence interval; HR, hazard ratio.

^a^ Adjusted for age, age-square, sex, TDI, education, CCI, and first 20 principal components of ancestry.

^b^ Adjusted for Model 1 and genetic risk categories

^c^ The trend test used the median value of each group instead of the original group.

**Supplementary Table 13**. Risk of incident Crohn's disease and ulcerative colitis by unweighted healthy lifestyle categories

| **Unweighted healthy lifestyle categories** | **Events/Person-years** | **Model 1 ^a^** | | **Model 2 ^b^** | |
| --- | --- | --- | --- | --- | --- |
|  |  | **HR (95% CI)** | ***P* value** | **HR (95% CI)** | ***P* value** |
| Crohn's disease |  |  |  |  |  |
| Favorable | 290/2,679,393 | 1 (Ref) |  | 1 (Ref) |  |
| Intermediate | 206/1,333,305 | 1.35 (1.13, 1.62) | <0.001 | 1.36 (1.13, 1.62) | <0.001 |
| Unfavorable | 201/875,841 | 1.94 (1.61, 2.33) | <0.001 | 1.94 (1.61, 2.33) | <0.001 |
| *p* value for trend ^c^ |  |  | <0.001 |  | <0.001 |
| Ulcerative colitis |  |  |  |  |  |
| Favorable | 254/1,176,187 | 1 (Ref) |  | 1 (Ref) |  |
| Intermediate | 513/1,925,357 | 1.34 (1.19, 1.51) | <0.001 | 1.34 (1.19, 1.51) | <0.001 |
| Unfavorable | 755/1,806,103 | 1.73 (1.53, 1.96) | <0.001 | 1.73 (1.53, 1.96) | <0.001 |
| *p* value for trend ^c^ |  |  | <0.001 |  | <0.001 |

CI indicates confidence interval; HR, hazard ratio.

^a^ Adjusted for age, age-square, sex, TDI, education, CCI, and first 20 principal components of ancestry.

^b^ Adjusted for Model 1 and genetic risk categories.

^c^ The trend test used the median value of each group instead of the original group.

**Supplementary Table 14**. Interactions of lifestyle factors and healthy lifestyle category with genetic risk.

| **Lifestyle Factors** | **Genetic risk of CD** | **Genetic risk of UC** |
| --- | --- | --- |
|  | ***P* value for interaction** | ***P* value for interaction** |
| Smoking status | 0.441 | 0.967 |
| Body mass index | 0.828 | 0.441 |
| Sleep duration | 0.021 | 0.459 |
| Diet | 0.266 | 0.485 |
| Physical activity | 0.800 | 0.536 |
| Healthy lifestyle category | 0.846 | 0.868 |
| Weighted healthy lifestyle category | 0.846 | 0.865 |

CD, Crohn's disease; UC, ulcerative colitis.

Adjusted for age, age-square, sex, TDI, education, CCI, and first 20 principal components of ancestry, and other healthy lifestyle factors.

**Supplementary Table 15**. Risk of incident Crohn’s disease by genetic and lifestyle risk with unweighted lifestyle score, and unweighted and weighted lifestyle score in multivariate imputed data, and after excluding participants with incomplete data of covariables

| **Subgroup** | **Unweighted lifestyle categories (n=429,515)** | | | **Unweighted lifestyle categories, multivariate imputed (n=429,515)** | | |
| --- | --- | --- | --- | --- | --- | --- |
|  | **Events/Person-years** | **HR (95% CI)** | ***P* value** | **Events/Person-years** | **HR (95% CI)** | ***P* value** |
| Low genetic risk |  |  |  |  |  |  |
| Favorable lifestyle | 36/537,238 | 1 (Ref) |  | 36/537,238 | 1 (Ref) |  |
| Intermediate lifestyle | 24/281,312 | 1.22 (0.73, 2.05) | 0.450 | 24/281,312 | 1.22 (0.73, 2.05) | 0.450 |
| Unfavorable lifestyle | 32/188,765 | 2.32 (1.44, 3.74) | 0.001 | 32/188,765 | 2.32 (1.44, 3.74) | 0.001 |
| Intermediate genetic risk | |  |  |  |  |  |
| Favorable lifestyle | 165/1,620,117 | 1.51 (1.06, 2.17) | 0.024 | 165/1,620,117 | 1.51 (1.06, 2.17) | 0.024 |
| Intermediate lifestyle | 124/836,165 | 2.11 (1.46, 3.06) | <0.001 | 124/836,165 | 2.11 (1.46, 3.06) | <0.001 |
| Unfavorable lifestyle | 119/566,705 | 2.85 (1.96, 4.15) | <0.001 | 119/566,705 | 2.85 (1.96, 4.15) | <0.001 |
| High genetic risk |  |  |  |  |  |  |
| Favorable lifestyle | 85/539,469 | 2.33 (1.58, 3.44) | <0.001 | 85/539,469 | 2.33 (1.58, 3.44) | <0.001 |
| Intermediate lifestyle | 61/278,415 | 3.12 (2.06, 4.71) | <0.001 | 61/278,415 | 3.12 (2.06, 4.71) | <0.001 |
| Unfavorable lifestyle | 61/188,296 | 4.40 (2.91, 6.66) | <0.001 | 61/188,296 | 4.40 (2.90, 6.66) | <0.001 |
| **Subgroup** | **Weighted lifestyle categories, multivariate imputed (n=429,515)** | | | **Participants with imcomplete data of covariables excluded (n=425,523)** | | |
|  | **Events/Person-years** | **HR (95% CI)** | ***P* value** | **Events/Person-years** | **HR (95% CI)** | ***P* value** |
| Low genetic risk |  |  |  |  |  |  |
| Favorable lifestyle | 36/537,238 | 1 (Ref) |  | 36/533,017 | 1 (Ref) |  |
| Intermediate lifestyle | 24/281,312 | 1.22 (0.73, 2.05) | 0.450 | 24/278,378 | 1.22 (0.73, 2.05) | 0.449 |
| Unfavorable lifestyle | 32/188,765 | 2.32 (1.44, 3.74) | 0.001 | 32/186,460 | 2.33 (1.44, 3.75) | 0.001 |
| Intermediate genetic risk | |  |  |  |  |  |
| Favorable lifestyle | 165/1,620,117 | 1.51 (1.06, 2.17) | 0.024 | 163/1,606,544 | 1.50 (1.04, 2.15) | 0.029 |
| Intermediate lifestyle | 124/836,165 | 2.11 (1.46, 3.06) | <0.001 | 124/827,234 | 2.11 (1.46, 3.07) | <0.001 |
| Unfavorable lifestyle | 119/566,705 | 2.85 (1.96, 4.15) | <0.001 | 119/560,155 | 2.85 (1.96, 4.15) | <0.001 |
| High genetic risk |  |  |  |  |  |  |
| Favorable lifestyle | 85/539,469 | 2.33 (1.58, 3.44) | <0.001 | 85/535,005 | 2.33 (1.58, 3.44) | <0.001 |
| Intermediate lifestyle | 61/278,415 | 3.12 (2.06, 4.71) | <0.001 | 60/275,735 | 3.06 (2.03, 4.64) | <0.001 |
| Unfavorable lifestyle | 61/188,296 | 4.40 (2.90, 6.66) | <0.001 | 59/186,279 | 4.25 (2.80, 6.46) | <0.001 |

CI indicates confidence interval; HR, hazard ratio.

**Supplementary Table 16**. Risk of incident ulcerative colitis by genetic and lifestyle risk with unweighted lifestyle score, and unweighted and weighted lifestyle score in multivariate imputed data, and after excluding participants with incomplete data of covariables

| **Subgroup** | **Unweighted lifestyle categories (n=430,384)** | | | **Unweighted lifestyle categories, multivariate imputed (n=430,384)** | | |
| --- | --- | --- | --- | --- | --- | --- |
|  | **Events/Person-years** | **HR (95% CI)** | ***P* value** | **Events/Person-years** | **HR (95% CI)** | ***P* value** |
| Low genetic risk |  |  |  |  |  |  |
| Favorable lifestyle | 90/538,340 | 1 (Ref) |  | 90/538,340 | 1 (Ref) |  |
| Intermediate lifestyle | 61/280,619 | 1.20 (0.87, 1.66) | 0.272 | 61/280,619 | 1.20 (0.87, 1.66) | 0.270 |
| Unfavorable lifestyle | 61/190,527 | 1.64 (1.18, 2.28) | 0.003 | 61/190,527 | 1.64 (1.19, 2.28) | 0.003 |
| Intermediate genetic risk | |  |  |  |  |  |
| Favorable lifestyle | 372/1,622,072 | 1.37 (1.09, 1.72) | 0.007 | 372/1,622,072 | 1.37 (1.09, 1.72) | 0.007 |
| Intermediate lifestyle | 284/838,688 | 1.86 (1.47, 2.37) | <0.001 | 284/838,688 | 1.87 (1.47, 2.37) | <0.001 |
| Unfavorable lifestyle | 254/564,710 | 2.31 (1.81, 2.94) | <0.001 | 254/564,710 | 2.31 (1.81, 2.94) | <0.001 |
| High genetic risk |  |  |  |  |  |  |
| Favorable lifestyle | 179/538,823 | 1.98 (1.54, 2.56) | <0.001 | 179/538,823 | 1.98 (1.54, 2.56) | <0.001 |
| Intermediate lifestyle | 137/278,418 | 2.72 (2.08, 3.55) | <0.001 | 137/278,418 | 2.72 (2.09, 3.55) | <0.001 |
| Unfavorable lifestyle | 138/190,140 | 3.73 (2.86, 4.88) | <0.001 | 138/190,140 | 3.74 (2.86, 4.88) | <0.001 |
| **Subgroup** | **Weighted lifestyle categories, multivariate imputed (n=430,384)** | | | **Participants with imcomplete data of covariables excluded (n=426,377)** | | |
|  | **Events/Person-years** | **HR (95% CI)** | ***P* value** | **Events/Person-years** | **HR (95% CI)** | ***P* value** |
| Low genetic risk |  |  |  |  |  |  |
| Favorable lifestyle | 84/538,409 | 1 (Ref) |  | 81/533,207 | 1 (Ref) |  |
| Intermediate lifestyle | 83/332,770 | 1.49 (1.10, 2.02) | 0.010 | 83/329,756 | 1.54 (1.14, 2.10) | 0.006 |
| Unfavorable lifestyle | 45/138,308 | 1.77 (1.23, 2.55) | 0.002 | 45/136,364 | 1.85 (1.28, 2.66) | 0.001 |
| Intermediate genetic risk | |  |  |  |  |  |
| Favorable lifestyle | 352/1,617,488 | 1.39 (1.10, 1.77) | 0.006 | 347/1,603,140 | 1.42 (1.12, 1.81) | 0.004 |
| Intermediate lifestyle | 353/996,460 | 2.11 (1.66, 2.68) | <0.001 | 348/987,264 | 2.16 (1.69, 2.75) | <0.001 |
| Unfavorable lifestyle | 205/411,522 | 2.71 (2.10, 3.50) | <0.001 | 205/406,626 | 2.82 (2.17, 3.65) | <0.001 |
| High genetic risk |  |  |  |  |  |  |
| Favorable lifestyle | 172/537,814 | 2.05 (1.58, 2.66) | <0.001 | 172/533,402 | 2.12 (1.63, 2.76) | <0.001 |
| Intermediate lifestyle | 169/331,001 | 3.06 (2.35, 3.97) | <0.001 | 165/327,558 | 3.09 (2.37, 4.04) | <0.001 |
| Unfavorable lifestyle | 113/138,566 | 4.45 (3.35, 5.91) | <0.001 | 110/137,246 | 4.49 (3.36, 5.99) | <0.001 |

CI indicates confidence interval; HR, hazard ratio.

**Supplementary Table 17**. Risk of Incident Crohn’s disease by genetic and lifestyle risk after excluding incident Crohn’s disease within 2, 3 years after baseline, after excluding baseline colorectal cancer, and additionally adjusted for depression

| **Subgroup** | **Incident CD within 2 years after baseline excluded (n=429,441)** | | | **Incident CD within 3 years after baseline excluded (n=429,386)** | | | **Prevalent colorectal cancer at baseline excluded (n=427,432)** | | |
| --- | --- | --- | --- | --- | --- | --- | --- | --- | --- |
|  | **Events/Person-years** | **HR (95% CI)** | ***P* value** | **Events/Person-years** | **HR (95% CI)** | ***P* value** | **Events/Person-years** | **HR (95% CI)** | ***P* value** |
| Low genetic risk |  |  |  |  |  |  |  |  |  |
| Favorable lifestyle | 31/537,230 | 1 (Ref) |  | 28/537,222 | 1 (Ref) |  | 35/534,975 | 1 (Ref) |  |
| Intermediate lifestyle | 22/281,309 | 1.30 (0.75, 2.25) | 0.341 | 22/281,309 | 1.46 (0.83, 2.55) | 0.188 | 24/279,998 | 1.26 (0.75, 2.12) | 0.387 |
| Unfavorable lifestyle | 27/188,757 | 2.30 (1.37, 3.86) | 0.002 | 24/188,750 | 2.30 (1.33, 3.97) | 0.003 | 31/187,851 | 2.32 (1.43, 3.77) | 0.001 |
| Intermediate genetic risk | |  |  |  |  |  |  |  |  |
| Favorable lifestyle | 150/1,620,096 | 1.60 (1.08, 2.35) | 0.018 | 137/1,620,062 | 1.62 (1.08, 2.43) | 0.021 | 162/1,613,772 | 1.53 (1.06, 2.20) | 0.023 |
| Intermediate lifestyle | 115/836,151 | 2.28 (1.53, 3.40) | <0.001 | 107/836,131 | 2.37 (1.56, 3.59) | <0.001 | 123/832,211 | 2.16 (1.48, 3.15) | <0.001 |
| Unfavorable lifestyle | 102/566,679 | 2.86 (1.91, 4.29) | <0.001 | 92/566,654 | 2.90 (1.90, 4.45) | <0.001 | 119/563,501 | 2.95 (2.01, 4.31) | <0.001 |
| High genetic risk |  |  |  |  |  |  |  |  |  |
| Favorable lifestyle | 75/539,454 | 2.38 (1.57, 3.62) | <0.001 | 68/539,436 | 2.39 (1.54, 3.72) | <0.001 | 84/537,556 | 2.36 (1.59, 3.51) | <0.001 |
| Intermediate lifestyle | 57/278,409 | 3.39 (2.19, 5.25) | <0.001 | 52/278,396 | 3.45 (2.18, 5.47) | <0.001 | 60/276,886 | 3.17 (2.08, 4.81) | <0.001 |
| Unfavorable lifestyle | 54/188,287 | 4.55 (2.92, 7.10) | <0.001 | 48/188,272 | 4.56 (2.85, 7.29) | <0.001 | 61/187,406 | 4.54 (2.99, 6.90) | <0.001 |
| **Subgroup** | **Additionally adjusted for depression (n=429,515)** | | |  | | |  | | |
|  | **Events/Person-years** | **HR (95% CI)** | ***P* value** |  |  |  |  |  |  |
| Low genetic risk |  |  |  |  |  |  |  |  |  |
| Favorable lifestyle | 36/537,238 | 1 (Ref) |  |  |  |  |  |  |  |
| Intermediate lifestyle | 24/281,312 | 1.22 (0.72, 2.04) | 0.460 |  |  |  |  |  |  |
| Unfavorable lifestyle | 32/188,765 | 2.28 (1.41, 3.69) | 0.001 |  |  |  |  |  |  |
| Intermediate genetic risk | |  |  |  |  |  |  |  |  |
| Favorable lifestyle | 165/1,620,117 | 1.52 (1.06, 2.17) | 0.024 |  |  |  |  |  |  |
| Intermediate lifestyle | 124/836,165 | 2.10 (1.45, 3.05) | <0.001 |  |  |  |  |  |  |
| Unfavorable lifestyle | 119/566,705 | 2.81 (1.93, 4.09) | <0.001 |  |  |  |  |  |  |
| High genetic risk |  |  |  |  |  |  |  |  |  |
| Favorable lifestyle | 85/539,469 | 2.33 (1.58, 3.44) | <0.001 |  |  |  |  |  |  |
| Intermediate lifestyle | 61/278,415 | 3.10 (2.05, 4.69) | <0.001 |  |  |  |  |  |  |
| Unfavorable lifestyle | 61/188,296 | 4.33 (2.86, 6.56) | <0.001 |  |  |  |  |  |  |

CI indicates confidence interval; HR, hazard ratio.

**Supplementary Table 18**. Risk of Incident ulcerative colitis by genetic and lifestyle risk after excluding incident ulcerative colitis within 2, 3 years after baseline, after excluding baseline colorectal cancer, and additionally adjusted for depression

| **Subgroup** | **Incident UC within 2 years after baseline excluded (n=430,247)** | | | **Incident UC within 3 years after baseline excluded (n=430,123)** | | | **Prevalent colorectal cancer at baseline excluded (n=428,298)** | | |
| --- | --- | --- | --- | --- | --- | --- | --- | --- | --- |
|  | **Events/Person-years** | **HR (95% CI)** | ***P* value** | **Events/Person-years** | **HR (95% CI)** | ***P* value** | **Events/Person-years** | **HR (95% CI)** | ***P* value** |
| Low genetic risk |  |  |  |  |  |  |  |  |  |
| Favorable lifestyle | 78/538,400 | 1 (Ref) |  | 71/538,383 | 1 (Ref) |  | 83/536,380 | 1 (Ref) |  |
| Intermediate lifestyle | 79/332,764 | 1.53 (1.12, 2.09) | 0.008 | 72/332,746 | 1.53 (1.10, 2.12) | 0.011 | 82/331,266 | 1.49 (1.10, 2.02) | 0.010 |
| Unfavorable lifestyle | 38/138,298 | 1.61 (1.09, 2.38) | 0.016 | 34/138,288 | 1.58 (1.05, 2.39) | 0.028 | 44/137,500 | 1.77 (1.23, 2.54) | 0.002 |
| Intermediate genetic risk | |  |  |  |  |  |  |  |  |
| Favorable lifestyle | 324/1,617,444 | 1.38 (1.08, 1.77) | 0.010 | 298/1,617,380 | 1.40 (1.08, 1.81) | 0.011 | 352/1,611,648 | 1.39 (1.10, 1.77) | 0.006 |
| Intermediate lifestyle | 325/996,418 | 2.10 (1.64, 2.69) | <0.001 | 296/996,343 | 2.10 (1.62, 2.72) | <0.001 | 349/991,329 | 2.11 (1.66, 2.68) | <0.001 |
| Unfavorable lifestyle | 183/411,491 | 2.61 (2.00, 3.41) | <0.001 | 167/411,451 | 2.61 (1.97, 3.45) | <0.001 | 205/409,210 | 2.70 (2.09, 3.49) | <0.001 |
| High genetic risk |  |  |  |  |  |  |  |  |  |
| Favorable lifestyle | 155/537,790 | 1.99 (1.51, 2.61) | <0.001 | 141/537,753 | 1.99 (1.49, 2.64) | <0.001 | 169/535,689 | 2.05 (1.58, 2.66) | <0.001 |
| Intermediate lifestyle | 152/330,975 | 2.96 (2.25, 3.89) | <0.001 | 144/330,955 | 3.08 (2.32, 4.09) | <0.001 | 168/329,118 | 3.06 (2.35, 3.97) | <0.001 |
| Unfavorable lifestyle | 105/138,553 | 4.46 (3.32, 6.00) | <0.001 | 92/138,522 | 4.28 (3.13, 5.85) | <0.001 | 113/137,854 | 4.43 (3.33, 5.89) | <0.001 |
| **Subgroup** | **Additionally adjusted for depression (n=430,384)** | | |  | | |  | | |
|  | **Events/Person-years** | **HR (95% CI)** | ***P* value** |  |  |  |  |  |  |
| Low genetic risk |  |  |  |  |  |  |  |  |  |
| Favorable lifestyle | 84/538,409 | 1 (Ref) |  |  |  |  |  |  |  |
| Intermediate lifestyle | 83/332,770 | 1.49 (1.10, 2.02) | 0.010 |  |  |  |  |  |  |
| Unfavorable lifestyle | 45/138,308 | 1.78 (1.23, 2.56) | 0.002 |  |  |  |  |  |  |
| Intermediate genetic risk | |  |  |  |  |  |  |  |  |
| Favorable lifestyle | 352/1,617,488 | 1.39 (1.10, 1.77) | 0.006 |  |  |  |  |  |  |
| Intermediate lifestyle | 353/996,460 | 2.11 (1.66, 2.68) | <0.001 |  |  |  |  |  |  |
| Unfavorable lifestyle | 205/411,522 | 2.72 (2.10, 3.51) | <0.001 |  |  |  |  |  |  |
| High genetic risk |  |  |  |  |  |  |  |  |  |
| Favorable lifestyle | 172/537,814 | 2.05 (1.58, 2.66) | <0.001 |  |  |  |  |  |  |
| Intermediate lifestyle | 169/331,001 | 3.05 (2.35, 3.97) | <0.001 |  |  |  |  |  |  |
| Unfavorable lifestyle | 113/138,566 | 4.46 (3.35, 5.93) | <0.001 |  |  |  |  |  |  |

CI indicates confidence interval; HR, hazard ratio.

**Supplementary Table 19**. Risk of Incident Crohn’s disease by genetic and lifestyle risk stratified by covariates

| **Subgroup** | **Age ≤ 60** | | **Age > 60** | | **Female** | | **Male** | | **College/University degree** | | **No college/university degree** | |
| --- | --- | --- | --- | --- | --- | --- | --- | --- | --- | --- | --- | --- |
|  | **HR (95% CI)** | ***P*** | **HR (95% CI)** | ***P*** | **HR (95% CI)** | ***P*** | **HR (95% CI)** | ***P*** | **HR (95% CI)** | ***P*** | **HR (95% CI)** | ***P*** |
| Low genetic risk |  |  |  |  |  |  |  |  |  |  |  |  |
| Favorable lifestyle | 1 (Ref) |  | 1 (Ref) |  | 1 (Ref) |  | 1 (Ref) |  | 1 (Ref) |  | 1 (Ref) |  |
| Intermediate lifestyle | 1.40 (0.66, 2.95) | 0.383 | 1.07 (0.52, 2.20) | 0.846 | 1.68 (0.89, 3.19) | 0.110 | 0.71 (0.29, 1.75) | 0.459 | 1.23 (0.42, 3.59) | 0.708 | 1.19 (0.66, 2.15) | 0.562 |
| Unfavorable lifestyle | 3.14 (1.61, 6.13) | 0.001 | 1.64 (0.81, 3.32) | 0.169 | 2.58 (1.34, 4.95) | 0.005 | 2.02 (0.99, 4.10) | 0.052 | 3.83 (1.51, 9.73) | 0.005 | 1.98 (1.13, 3.46) | 0.016 |
| Intermediate genetic risk | |  |  |  |  |  |  |  |  |  |  |  |
| Favorable lifestyle | 2.04 (1.20, 3.46) | 0.008 | 1.09 (0.66, 1.81) | 0.723 | 1.59 (0.99, 2.54) | 0.054 | 1.43 (0.81, 2.50) | 0.217 | 1.96 (1.00, 3.82) | 0.050 | 1.34 (0.87, 2.06) | 0.178 |
| Intermediate lifestyle | 2.81 (1.63, 4.83) | <0.001 | 1.56 (0.93, 2.61) | 0.093 | 2.23 (1.36, 3.64) | 0.001 | 1.93 (1.09, 3.42) | 0.023 | 3.07 (1.53, 6.17) | 0.002 | 1.81 (1.17, 2.81) | 0.008 |
| Unfavorable lifestyle | 3.79 (2.19, 6.55) | <0.001 | 2.10 (1.24, 3.55) | 0.006 | 3.40 (2.07, 5.59) | <0.001 | 2.28 (1.28, 4.05) | 0.005 | 2.71 (1.24, 5.92) | 0.013 | 2.78 (1.80, 4.28) | <0.001 |
| High genetic risk |  |  |  |  |  |  |  |  |  |  |  |  |
| Favorable lifestyle | 2.85 (1.61, 5.04) | <0.001 | 1.92 (1.12, 3.29) | 0.018 | 2.41 (1.45, 4.00) | 0.001 | 2.23 (1.22, 4.10) | 0.010 | 2.48 (1.19, 5.14) | 0.015 | 2.27 (1.43, 3.61) | <0.001 |
| Intermediate lifestyle | 4.78 (2.68, 8.52) | <0.001 | 1.80 (0.97, 3.35) | 0.064 | 2.83 (1.61, 4.99) | <0.001 | 3.33 (1.81, 6.15) | <0.001 | 2.65 (1.12, 6.24) | 0.026 | 3.18 (1.98, 5.11) | <0.001 |
| Unfavorable lifestyle | 5.92 (3.27, 10.71) | <0.001 | 3.18 (1.76, 5.75) | <0.001 | 5.82 (3.40, 9.97) | <0.001 | 3.06 (1.60, 5.85) | 0.001 | 5.23 (2.22, 12.34) | <0.001 | 4.11 (2.55, 6.61) | <0.001 |
|  | **TDI_T1** | | **TDT_T2** | | **TDI_T3** | | **Charlson comorbidities index = 0** | | **Charlson comorbidities index > 0** | |  |  |
|  | **HR (95% CI)** | ***P*** | **HR (95% CI)** | ***P*** | **HR (95% CI)** | ***P*** | **HR (95% CI)** | ***P*** | **HR (95% CI)** | ***P*** |  |  |
| Low genetic risk |  |  |  |  |  |  |  |  |  |  |  |  |
| Favorable lifestyle | 1 (Ref) |  | 1 (Ref) |  | 1 (Ref) |  | 1 (Ref) |  | 1 (Ref) |  |  |  |
| Intermediate lifestyle | 0.57 (0.19, 1.73) | 0.324 | 1.09 (0.40, 2.96) | 0.860 | 2.13 (0.95, 4.80) | 0.068 | 1.39 (0.79, 2.44) | 0.251 | 0.60 (0.15, 2.32) | 0.458 |  |  |
| Unfavorable lifestyle | 1.25 (0.45, 3.44) | 0.669 | 2.63 (1.09, 6.35) | 0.032 | 3.32 (1.53, 7.21) | 0.002 | 2.61 (1.54, 4.45) | <0.001 | 1.33 (0.44, 3.97) | 0.613 |  |  |
| Intermediate genetic risk | |  |  |  |  |  |  |  |  |  |  |  |
| Favorable lifestyle | 1.17 (0.66, 2.07) | 0.599 | 1.60 (0.84, 3.07) | 0.154 | 1.93 (0.99, 3.77) | 0.055 | 1.57 (1.05, 2.34) | 0.028 | 1.30 (0.57, 2.97) | 0.536 |  |  |
| Intermediate lifestyle | 1.53 (0.83, 2.82) | 0.177 | 2.99 (1.55, 5.75) | 0.001 | 2.18 (1.09, 4.35) | 0.027 | 2.33 (1.54, 3.51) | <0.001 | 1.27 (0.53, 3.02) | 0.592 |  |  |
| Unfavorable lifestyle | 2.13 (1.12, 4.03) | 0.021 | 3.75 (1.91, 7.34) | <0.001 | 3.28 (1.66, 6.46) | 0.001 | 2.89 (1.89, 4.41) | <0.001 | 2.33 (1.02, 5.32) | 0.044 |  |  |
| High genetic risk |  |  |  |  |  |  |  |  |  |  |  |  |
| Favorable lifestyle | 1.86 (0.99, 3.49) | 0.052 | 2.43 (1.20, 4.89) | 0.013 | 2.92 (1.43, 5.97) | 0.003 | 2.28 (1.48, 3.53) | <0.001 | 2.48 (1.03, 5.93) | 0.042 |  |  |
| Intermediate lifestyle | 2.72 (1.38, 5.35) | 0.004 | 2.56 (1.16, 5.65) | 0.02 | 4.26 (2.07, 8.78) | <0.001 | 2.84 (1.77, 4.55) | <0.001 | 3.73 (1.55, 8.94) | 0.003 |  |  |
| Unfavorable lifestyle | 2.96 (1.38, 6.33) | 0.005 | 5.32 (2.51, 11.29) | <0.001 | 5.66 (2.76, 11.58) | <0.001 | 4.32 (2.69, 6.93) | <0.001 | 3.94 (1.63, 9.51) | 0.002 |  |  |

CI indicates confidence interval; HR, hazard ratio; TDI, Townsend deprivation index.

**Supplementary Table 20**. Risk of Incident ulcerative colitis by genetic and lifestyle risk stratified by covariates

| **Subgroup** | **Age ≤ 60** | | **Age > 60** | | **Female** | | **Male** | | **College/University degree** | | **No college/university degree** | |
| --- | --- | --- | --- | --- | --- | --- | --- | --- | --- | --- | --- | --- |
|  | **HR (95% CI)** | ***P*** | **HR (95% CI)** | ***P*** | **HR (95% CI)** | ***P*** | **HR (95% CI)** | ***P*** | **HR (95% CI)** | ***P*** | **HR (95% CI)** | ***P*** |
| Low genetic risk |  |  |  |  |  |  |  |  |  |  |  |  |
| Favorable lifestyle | 1 (Ref) |  | 1 (Ref) |  | 1 (Ref) |  | 1 (Ref) |  | 1 (Ref) |  | 1 (Ref) |  |
| Intermediate lifestyle | 1.53 (1.01, 2.31) | 0.045 | 1.43 (0.91, 2.24) | 0.118 | 1.73 (1.12, 2.67) | 0.013 | 1.28 (0.83, 1.96) | 0.260 | 1.31 (0.72, 2.37) | 0.371 | 1.57 (1.10, 2.24) | 0.013 |
| Unfavorable lifestyle | 1.55 (0.92, 2.61) | 0.102 | 1.97 (1.18, 3.30) | 0.009 | 1.94 (1.10, 3.43) | 0.022 | 1.60 (0.99, 2.57) | 0.055 | 2.00 (0.90, 4.42) | 0.088 | 1.75 (1.16, 2.65) | 0.008 |
| Intermediate genetic risk | |  |  |  |  |  |  |  |  |  |  |  |
| Favorable lifestyle | 1.40 (1.02, 1.92) | 0.036 | 1.39 (0.97, 2.00) | 0.077 | 1.54 (1.10, 2.15) | 0.013 | 1.26 (0.90, 1.77) | 0.173 | 1.33 (0.87, 2.05) | 0.189 | 1.42 (1.07, 1.89) | 0.015 |
| Intermediate lifestyle | 2.26 (1.65, 3.11) | <0.001 | 1.93 (1.35, 2.77) | <0.001 | 2.46 (1.75, 3.46) | <0.001 | 1.81 (1.29, 2.52) | 0.001 | 2.07 (1.34, 3.20) | 0.001 | 2.15 (1.61, 2.85) | <0.001 |
| Unfavorable lifestyle | 2.85 (2.03, 4.02) | <0.001 | 2.51 (1.70, 3.70) | <0.001 | 3.15 (2.16, 4.60) | <0.001 | 2.35 (1.66, 3.34) | <0.001 | 2.38 (1.40, 4.05) | 0.001 | 2.81 (2.09, 3.79) | <0.001 |
| High genetic risk |  |  |  |  |  |  |  |  |  |  |  |  |
| Favorable lifestyle | 2.18 (1.55, 3.07) | <0.001 | 1.87 (1.25, 2.81) | 0.002 | 2.29 (1.58, 3.30) | <0.001 | 1.82 (1.25, 2.64) | 0.002 | 1.85 (1.15, 2.99) | 0.011 | 2.14 (1.57, 2.92) | <0.001 |
| Intermediate lifestyle | 3.37 (2.37, 4.78) | <0.001 | 2.07 (1.82, 4.01) | <0.001 | 3.34 (2.28, 4.87) | <0.001 | 2.78 (1.93, 4.00) | <0.001 | 2.96 (1.82, 4.83) | <0.001 | 3.11 (2.28, 4.25) | <0.001 |
| Unfavorable lifestyle | 4.51 (3.07, 6.62) | <0.001 | 4.32 (2.82, 6.61) | <0.001 | 4.73 (3.05, 7.32) | <0.001 | 4.06 (2.78, 5.94) | <0.001 | 4.87 (2.73, 8.67) | <0.001 | 4.41 (3.17, 6.14) | <0.001 |
|  | **TDI_T1** | | **TDT_T2** | | **TDI_T3** | | **Charlson comorbidities index = 0** | | **Charlson comorbidities index > 0** | |  |  |
|  | **HR (95% CI)** | ***P*** | **HR (95% CI)** | ***P*** | **HR (95% CI)** | ***P*** | **HR (95% CI)** | ***P*** | **HR (95% CI)** | ***P*** |  |  |
| Low genetic risk |  |  |  |  |  |  |  |  |  |  |  |  |
| Favorable lifestyle | 1 (Ref) |  | 1 (Ref) |  | 1 (Ref) |  | 1 (Ref) |  | 1 (Ref) |  |  |  |
| Intermediate lifestyle | 1.37 (0.74, 2.53) | 0.315 | 1.44 (0.86, 2.42) | 0.164 | 1.54 (0.95, 2.50) | 0.078 | 1.41 (1.01, 1.98) | 0.044 | 1.81 (0.89, 3.67) | 0.100 |  |  |
| Unfavorable lifestyle | 1.69 (0.76, 3.77) | 0.200 | 2.25 (1.24, 4.08) | 0.008 | 1.45 (0.82, 2.56) | 0.201 | 1.51 (0.98, 2.34) | 0.061 | 2.48 (1.19, 5.18) | 0.016 |  |  |
| Intermediate genetic risk | |  |  |  |  |  |  |  |  |  |  |  |
| Favorable lifestyle | 1.71 (1.10, 2.64) | 0.017 | 1.42 (0.95, 2.11) | 0.085 | 1.12 (0.75, 1.68) | 0.585 | 1.39 (1.07, 1.80) | 0.012 | 1.42 (0.78, 2.59) | 0.257 |  |  |
| Intermediate lifestyle | 2.58 (1.65, 4.02) | <0.001 | 1.93 (1.29, 2.90) | 0.001 | 1.93 (1.30, 2.86) | 0.001 | 2.12 (1.63, 2.75) | <0.001 | 2.08 (1.14, 3.77) | 0.016 |  |  |
| Unfavorable lifestyle | 2.85 (1.72, 4.74) | <0.001 | 2.39 (1.53, 3.74) | <0001 | 2.72 (1.81, 4.08) | <0.001 | 2.63 (1.98, 3.50) | <0.001 | 2.86 (1.56, 5.25) | 0.001 |  |  |
| High genetic risk |  |  |  |  |  |  |  |  |  |  |  |  |
| Favorable lifestyle | 2.47 (1.54, 3.97) | <0.001 | 2.04 (1.32, 3.16) | 0.001 | 1.70 (1.08, 2.67) | 0.022 | 2.01 (1.51, 2.68) | <0.001 | 2.24 (1.17, 4.30) | 0.015 |  |  |
| Intermediate lifestyle | 3.21 (1.95, 5.29) | <0.001 | 3.40 (2.20, 5.25) | <0.001 | 2.61 (1.69, 4.05) | <0.001 | 3.00 (2.25, 4.01) | <0.001 | 3.23 (1.70, 6.12) | <0.001 |  |  |
| Unfavorable lifestyle | 6.84 (4.03, 11.60) | <0.001 | 2.84 (1.64, 4.94) | <0.001 | 4.29 (2.76, 6.68) | <0.001 | 4.52 (3.29, 6.20) | <0.001 | 4.18 (2.15, 8.15) | <0.001 |  |  |

CI indicates confidence interval; HR, hazard ratio; TDI, Townsend deprivation index.

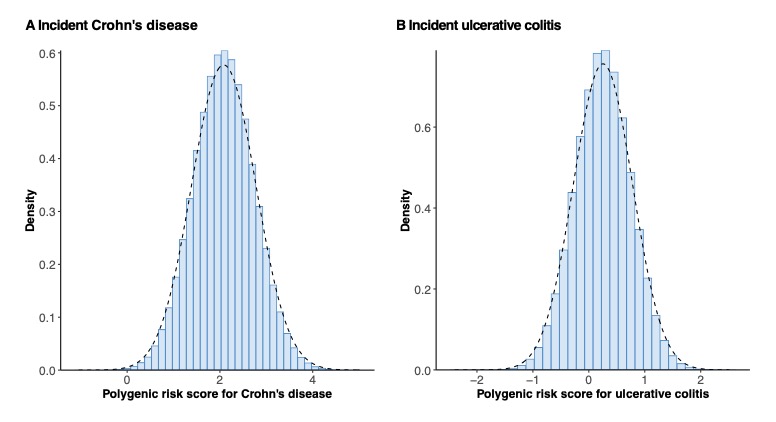

**Supplementary Figure 1**. Distribution of the polygenic risk score for Crohn’s disease (A) and ulcerative colitis (B).
